# The impact of genomic structural variation on the transcriptome, chromatin, and proteome in the human brain

**DOI:** 10.1101/2021.02.25.21252245

**Authors:** Ricardo A. Vialle, Katia de Paiva Lopes, David A. Bennett, John F. Crary, Towfique Raj

## Abstract

Structural variants (SVs), defined as any genomic rearrangements of 50 or more bp, are an important source of genetic diversity and have been linked to many diseases. However, their contribution to molecular traits in the brain and impact on neurodegenerative diseases remains unknown. Here, we report 170,996 SVs which were constructed using 1,760 short-read whole genomes from aging and Alzheimer’s disease subjects. We quantified the impact of *cis*-acting SVs on several molecular traits including histone modification, gene expression, mRNA splicing, and protein abundance in post-mortem brain tissues. More than 3,800 genes were associated with at least one molecular phenotype, and 712 (18%) with more than one phenotype, with a significant positive correlation in the direction of effect between RNA, histone peaks, and protein levels. SV associations with RNA and protein levels shared the same direction of effect in more than 87% of SV-gene pairs. We found reproducibility of SV-eQTLs across three groups of samples and multiple brain regions ranging from 81 to 98%, including the innate immune system related genes *ERAP2* and *GBP3*. Additionally, associations of SVs with progressive supranuclear palsy, an amyloid-independent primary tauopathy, identified previously known and novel SVs at the 17q.21.31 *MAPT* locus and several other novel suggestive associations. Our study provides a comprehensive view of the mechanisms linking structural variation to gene regulation and provides a valuable resource for understanding the functional impact of SVs in the aged human brain.

## BACKGROUND

Structural variants (SVs) are defined as genomic rearrangements ranging from fifty to thousands of base pairs^1^. These rearrangements can be classified as unbalanced (e.g., deletions, duplications, and insertions), balanced (e.g., inversions and translocations), or any complex combination of SV classes. SVs are widespread in the human genome and provide an important source of variation during evolution^2,3^. Current estimates suggest that a human genome may harbor up to 27,000 SVs compared to the reference genome^4^. In contrast to single-nucleotide polymorphisms (SNPs) and small indels, SVs can affect a higher fraction of the human genome^5^, suggesting that they may have more significant, or at least similar, consequences for phenotypic variation and evolution^2,3^.

The occurrence of an SV is commonly mediated by distal or proximal sequence homology around breakpoints, usually surrounded by mobile element insertions (MEIs), causing non-allelic homologous recombination (NAHR)^6^, or by DNA double-strand repair mechanisms such as non-homologous end-joining (NHEJ)^7,8^. Detecting these mutational signatures is challenging, and its correct classification depends on proper breakpoint resolution. Compared to low-resolution microarray technologies, whole-genome sequencing (WGS) data improves SV discovery by detecting more classes of variation, achieving higher SV size coverage and single base pair breakpoint resolution. With the increasing number of short-read WGS data produced, the number of genome-wide studies of SVs have been escalating in the past few years, jumping from 2,504 human genomes analyzed in the 1000 Genome Project^1^ to 14,891 in GnomAD^9^ and 17,795 in NHGRI Centers for Common Disease^10^. Nevertheless, we are still far from a complete and comprehensive population-scale human structural variation catalog.

Most studies on the impact of SVs so far have been restricted to non-brain tissues or to mRNA expression level only^11–13^. Large cohort studies, like the GTEx consortium, have already started mapping the impact of common and rare SVs on RNA expression^14^. The contribution of SVs in brain-related disorders and traits such as schizophrenia^15–17^, autism spectrum disorder (ASD)^18–20^, and cognition^21,22^ is notable. Genes expressed in brain tissues have complex features, with one of the highest expression levels and transcriptome complexity^23^, the longest introns^24^, more alternatively spliced intron clusters^12^, along with complex regulatory architecture^25^, making them especially vulnerable to SVs of all types. The effects of genetic variants can be modulated at different levels of gene regulation^11–13^. Therefore, identifying the impact of SVs on different molecular phenotypes in the brain is crucial to understanding their functional outcome and role in diseases. Here, we discovered SVs in 1,760 donors and, by integrating multi-omics data sets consisted of histone acetylation (H3K9ac, ChIP-seq), RNA (RNA-seq), and proteomics (TMT-Mass Spectrometry) measured in brain tissues for subsets of the same donors, we mapped the impact of common SVs into multiple molecular phenotypes. We measured the main SVs features associated with each phenotype and the propagation of effects through the regulatory cascade (**Figure 1**). We also identified pathogenic SVs related to neurodegenerative diseases and the impact of rare SVs on RNA and protein levels.

**Figure 1.**
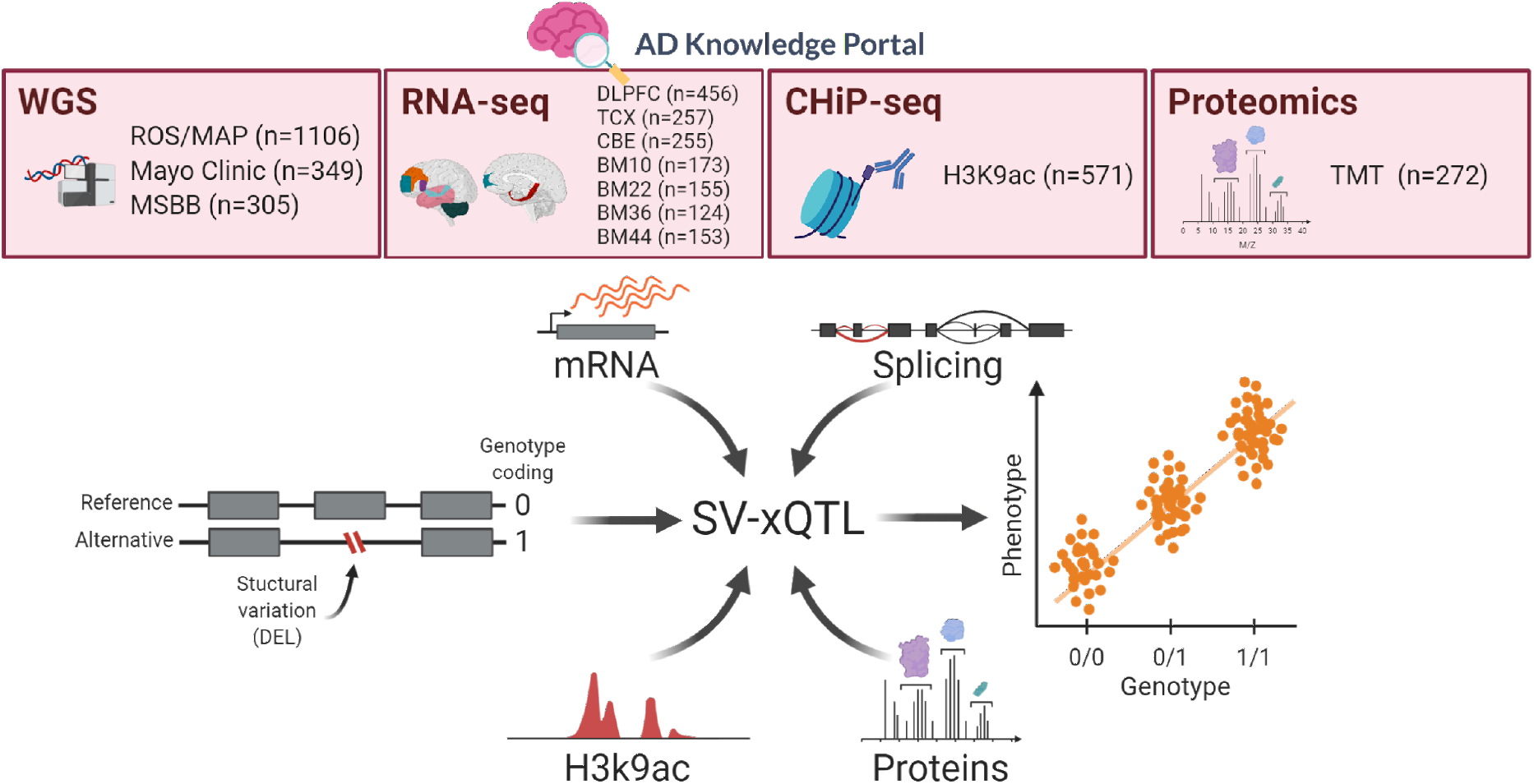
Study overview. The datasets used in this study have been made available to the research community through the Accelerating Medicines Partnership in Alzheimer’s Disease (AMP-AD) Knowledge Portal. Whole-genome sequencing and RNA-seq datasets are available from four aging and Alzheimer’s disease cohorts: Religious Orders Study (ROS) and Memory and Aging Project (MAP), Mayo Clinic, and Mount Sinai Brain Bank (MSBB). RNA-seq data for ROS/MAP are from the dorsolateral prefrontal cortex (DLPFC). RNA-seq data from MSBB are from four brain regions: BM10 = Brodmann area 10 (part of the frontopolar prefrontal cortex), BM22 = Brodmann area 22 (part of the superior temporal gyrus), BM36 = Brodmann area 36 (part of the fusiform gyrus), and BM44 = Brodmann area 44 (opercular part of the inferior frontal gyrus). RNA-seq from Mayo Clinic are from TCX = temporal cortex, CBE = cerebellum. The ChIP-seq (Histone 3 Lysine 9 acetylation, H3K9Ac) and proteomics data (Tandem mass tag, TMT) are from ROS/MAP cohorts. The post-QC sample sizes are shown next to each dataset. eQTL analyses were performed in all datasets; sQTL, haQTL, and pQTL were only performed with ROS/MAP data.

## RESULTS

### Structural variation discovery and quality assessment

We analyzed 1,881 human samples with WGS data generated from four cohorts (ROS/MAP, MSBB, and Mayo Clinic). To identify SVs in each group, we run a combination of seven different tools (see Methods) to capture the main classes of variation, including deletions (DEL), duplications (DUP), insertions (INS), inversions (INV), mobile element insertions (MEI), and complex rearrangements (CPX). These variants were further merged and genotyped at the group level (**Supplementary Figure S1**). After pre- and post-discovery quality control (QC. **Supplementary Table S1, Supplementary Figure S2**), a total of 170,966 ‘high-confidence’ SVs were identified in 1,760 samples that were used for all subsequent downstream analyses (**Figure 2a**). As expected, more SVs were detected in the ROS/MAP cohorts due to the larger sample size (n=1,106). More SVs were detected in MSBB compared to Mayo, due to ancestry differences^1,9^ as the Mayo Clinic group is composed of European ancestry individuals only, while MSBB has more diverse populations, including individuals of African and Admixed American ancestry (**Supplementary Figure S3**). Most SVs were small (median size of 280 bp), comprised by mostly deletions and insertions, with a decreasing frequency as the variants increased in size and with a high number of *Alu*, SVA, and LINE1 mobile element insertions identified (**Figure 2b**).

**Figure 2-.**
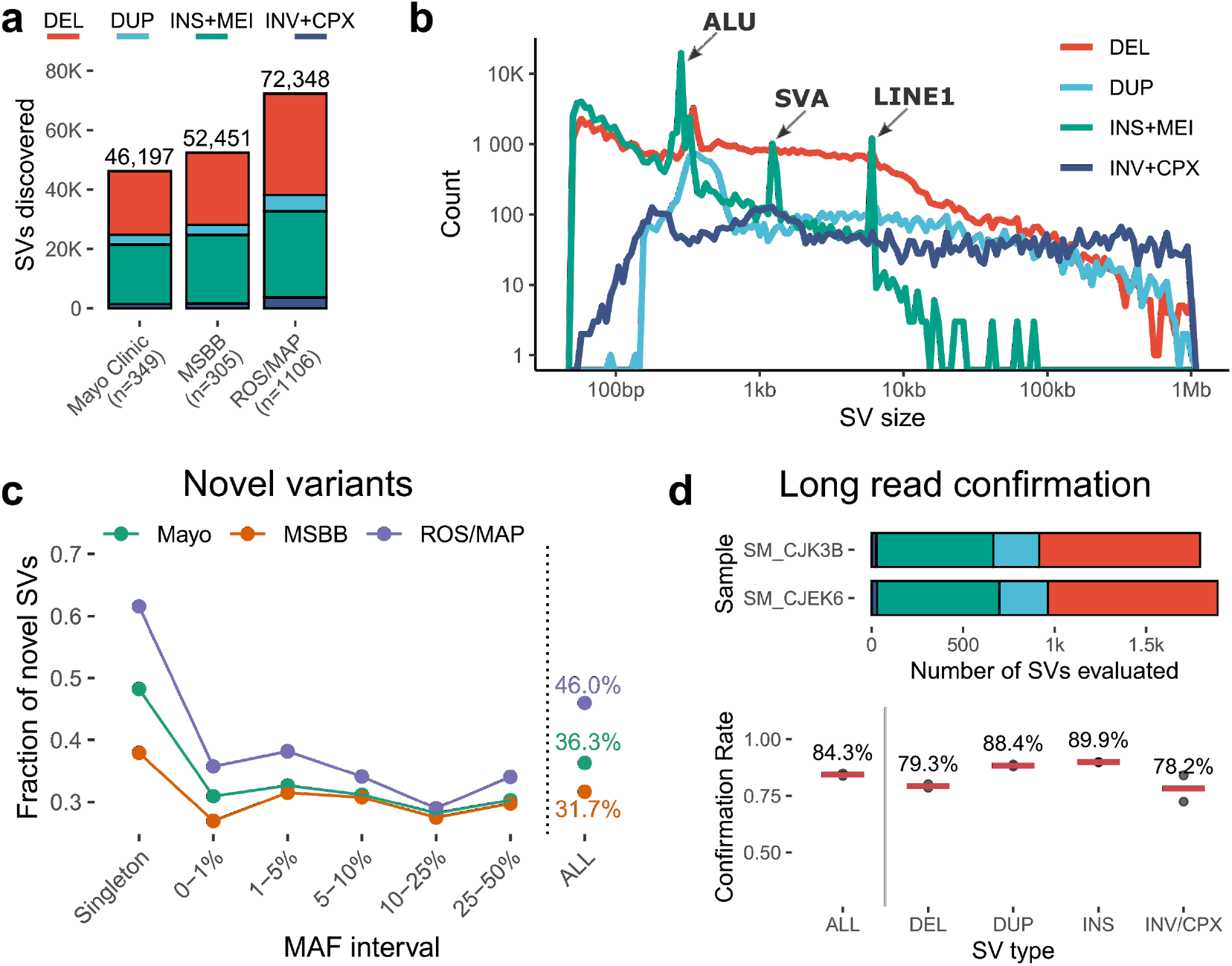
Summary of SV calls across cohorts. **a**, Total number of SVs identified within each cohort (ROS/MAP, Mayo Clinic, MSBB), colored by main SV types (DEL, DUP, INS+MEI, and INV+CPX). **b**, SV size distribution per SV type with x-axis and y-axis shown in log_10_ scale. **c**, Proportion of novel SVs found in each cohort stratified by minor allele frequency (MAF) spectrum. SVs were considered novel if not found in dbVar, Centers for Common Disease Genomics (CCDG), Database of Genomic Variants (DGV), Deciphering Developmental Disorders (DDD), GnomAD-SV, and the 1000 Genomes Project. **d**, Barplot showing samples sequenced using PacBio’s long-read WGS and number of SVs from short-reads evaluated for replication, plot below shows the confirmation rates for each sample (dots) measured using *VaPoR* and stratified by each SV class. Horizontal bars represent the median of both samples.

To assess the quality of SVs discovered, we first measured the reproducibility of our calls compared to other large datasets, including dbVar^26^, Centers for Common Disease Genomics (CCDG)^10^, Database of Genomic Variants (DGV)^27^, Deciphering Developmental Disorders (DDD)^28^, GnomAD-SV^9^, and 1000 Genomes Project^1^. We found about 30% of novel SVs and, as expected, the highest proportion of these SVs were discovered as singletons (**Figure 2c**). Further, allele frequency comparisons to SVs in common with the 1000 Genomes Project and GnomAD-SV showed high overall reproducibility with R^2^ equal to 0.84 and 0.67, respectively (**Supplementary Figure S4**). We also observed that about 75% of SVs were in Hardy–Weinberg equilibrium depending on the study (**Supplementary Figure S5**). In addition, we generated long-read WGS with PacBio for two ROS/MAP samples. We performed *in silico* confirmation of 4,581 SVs identified with short-reads and accessed a confirmation of 84.3% of them (**Figure 2d**). Together, these analyses provided sufficient evidence for the quality of the SVs discovered across all samples.

In accordance with previous studies^1,9,14,29,30^, a substantial proportion of SVs detected were rare (71%, minor allele frequency (MAF) < 0.05). More than 30% of SVs were observed in only one individual (i.e., singletons) (**Supplementary Figure S6a**). Additionally, by overlapping SVs with genomic annotations, we observed that singletons were more likely to occur in coding and regulatory regions compared to all other SVs (**Supplementary Figure S6b**). Moreover, constrained genes, such as morbid genes, loss-of-function (LoF) intolerant, and haploinsufficient genes, were more likely to be disrupted by singletons and ultra-rare SVs, reflecting the effects of purifying selection (**Supplementary Figure S6c-e**). These analyses demonstrate that the structural variants found here conform with principles of population genetics and highlight the importance of large sample sizes to improve the characterization of rare and pathogenic variants.

### Effects of SVs on gene expression

We performed associations of common SVs with gene expression in *cis* for the available brain regions (**Figure 3a**). The number of associations was highly correlated with the sample size (Pearson’s *r* 0.98, *P-*value 5 × 10^−5^). 98% of shared SV-eQTL showed the same direction of effect (β) (**Supplementary Figure S7**). The reproducibility of SV-eQTL across studies, as measured by Storey’s π1, showed substantial sharing of effects on brain gene expression. The highest reproducibility was observed within regions from the same studies, as a consequence of repeated donors (77,1% and 86.7% of donors from Mayo Clinic and MSBB, respectively, had RNA-seq for more than one brain region). However, regional effects were also observed when comparing different studies, for example, TCX and DLPFC shared more effects than DLPFC and CBE (0.93 and 0.81, respectively) (**Figure 3b**), suggesting some degree of regional specificity.

**Figure 3.**
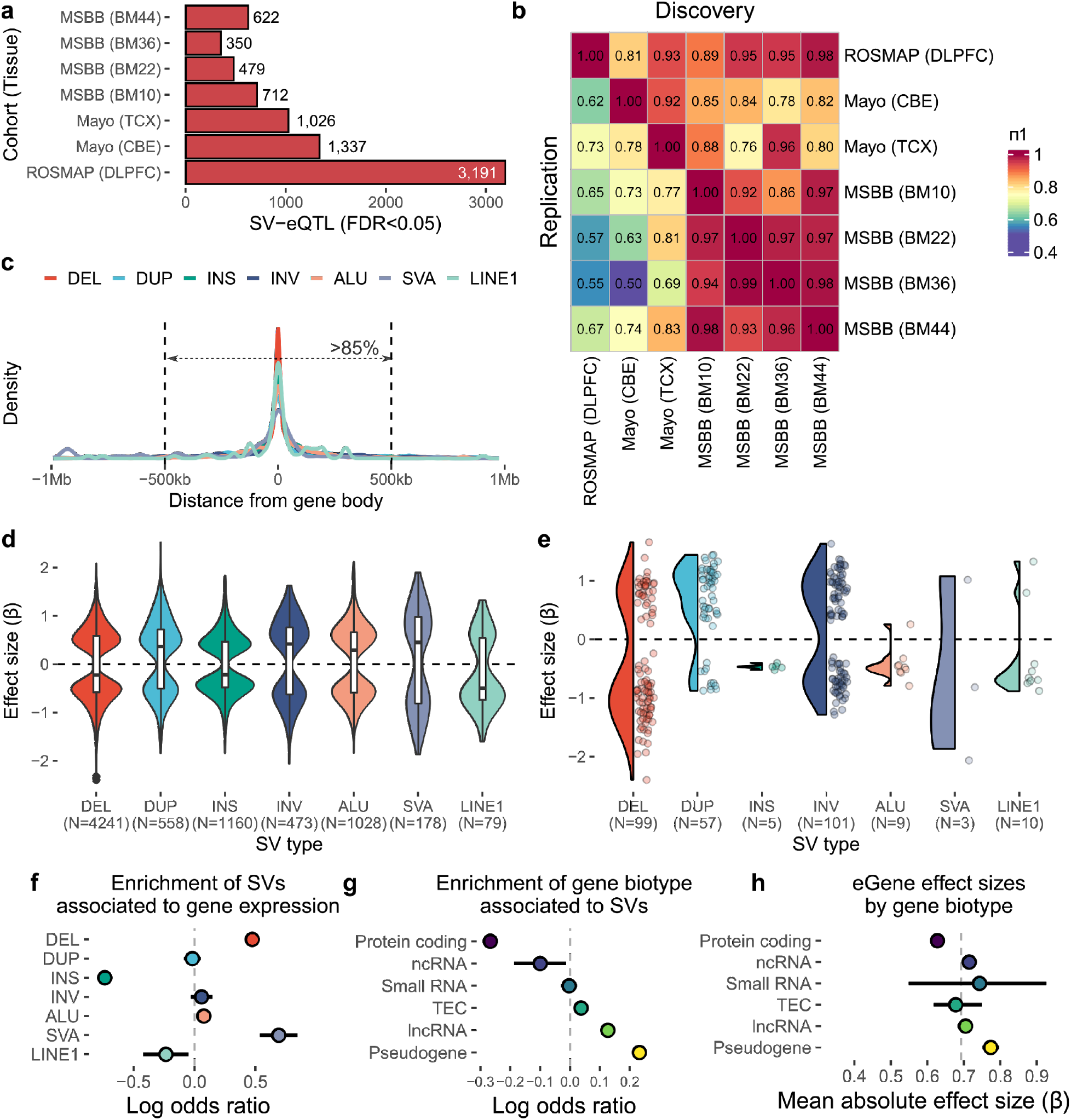
Properties of SV-eQTLs. **a**, Total number of significant SV-eQTLs (FDR < 0.05) identified within each cohort (ROS/MAP, Mayo Clinic, MSBB) in each brain region. **b**, SV-eQTL sharing across different groups and regions measured by π1 from qvalue R package. Columns represent the discovery sets while rows represent the replication set. **c**, Proportion of significant SVs by distance to the gene affected. More than 85% of significant associations across all SV types were within 500 kb distance of the gene. **d**, Distribution of effect sizes for each SV type. **e**, Distribution of effect sizes for SVs that overlap exonic regions of the associated gene. **f**, Log odds ratio of SV being associated with gene expression changes (i.e., being an SV-eQTL). Lines indicate 95% Wald confidence intervals. **g**, Log odds ratio of a gene being significantly associated stratified by gene biotype, lines indicate 95% Wald confidence intervals. **h**, Average absolute effect sizes of each eGene stratified by gene biotype, lines represent 95% confidence intervals (n = 1000 bootstraps).

More than 85% of the SVs associated with gene expression were within 500kb distance from the gene body (**Figure 3c**). The direction of effects (β) of SV-eQTLs was usually distributed in both directions (**Figure 3d**), except when the SVs were overlapping the exons (3.6%), in these cases, we observed differences likely caused by a disruption in the coding region; for instance, deletions or exon-disrupting MEIs causing decreased expression, while exon duplications would cause increased expression (**Figure 3e**). DELs and SVA transposons were more likely to be associated with changes in expression, while INS were less likely (**Figure 3f**). Pseudogenes, long non-coding RNAs (lncRNAs), and TEC (To be Experimentally Confirmed) were significantly more likely to be associated with SVs, while protein-coding and ncRNAs were less likely (**Figure 3g**). Additionally, the overall effect sizes of associations with pseudogenes were higher than protein-coding genes (**Figure 3h**). Such differences support evidence that less constrained genes (such as pseudogenes) are more likely to be eGenes in agreement with results previously observed for SV and SNV eQTLs^30,31^.

### Mapping of SVs that affect the gene-regulatory cascade

We mapped associations of 25,421 SVs with MAF ≥ 0.01 in the ROS/MAP cohorts to four different molecular phenotypes in the DLPFC. These molecular phenotypes included gene expression for 15,582 genes (n=456), 110,092 splicing junctions proportions measured by “percent spliced in” values (PSI) (n=505), histone acetylation (H3K9ac) peaks (n=571), and proteomic data for 7,960 proteins (n=272). We refer to these analyses as SV-xQTL, in which we map differences in measurements of each molecular phenotype associated with specific SV’s (**Figure 1**). Therefore, each SV-xQTL is an SV-phenotype pair (i.e., SV-eQTL, SV-sQTL, SV-haQTL, or SV-pQTL). All phenotype measurements were adjusted prior to associations to account for known (e.g., sex and ancestry principal components) and unknown covariates, determined either with PEER (probabilistic estimation of expression residuals) or PCA (principal component analysis) (see Methods). This, identified 3,191 SV-eQTL, 2,866 SV-sQTL, 399 SV-pQTL, and 1,454 SV-haQTL (FDR < 0.05) (**Figure 4a, Supplementary Figure S8**).

**Figure 4.**
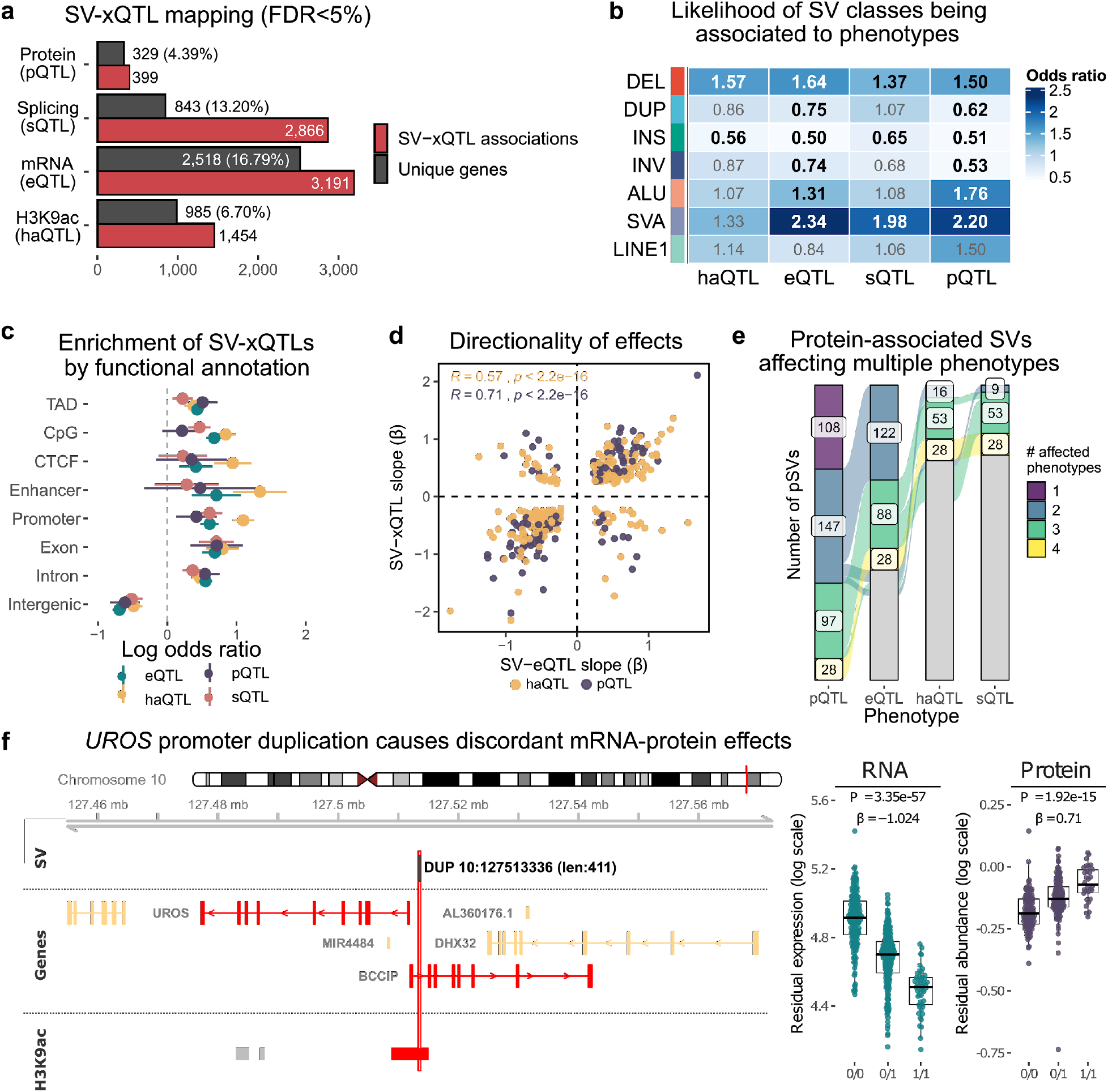
Impact of SVs on the gene-regulatory cascade. **a**, Total number of SV-xQTLs (FDR < 0.05) identified in ROS/MAP. Red bars show the number of lead per phenotype associations measured for each SV class separately, while gray bars show the total number of unique genes associated independently of SV classes. Percentages shown in the gene bars refer to the total number of genes tested for each phenotype. **b**, Heatmap showing the odds ratio of each SV class being associated with changes in each phenotype (i.e., being an SV-xQTL). Odds are measured against all lead SVs per phenotype, including non-significant. Numbers in bold represent *P*-value < 5% (Wald’s test). **c**, Enrichment of xSVs (i.e., SVs significantly associated to some phenotype) by functional annotation. Values are given as the log odds ratio of an xSV being overlapping a given genomic feature compared to all SVs tested for each molecular phenotype separately. Lines indicate 95% Wald confidence intervals. **d**, Slope correlation of SV-haQTL and SV-pQTL effect sizes (y-axis) compared to SV-eQTL effect sizes (x-axis). Pearson correlations and respective *P*-values are shown for each pair. **e**, SVs associated with proteins (380 pSVs, first bar) that are also associated with different molecular phenotypes (indicated at respective columns). Each color represents pSVs where the same SV-gene pair is significantly associated with a different number of phenotypes, from 1 (only at protein level) to 4 (all molecular phenotypes). **f**, Example of discordant effect between RNA and protein caused by a 411 bp duplication overlapping an H3K9ac peak upstream of the *UROS*. In the locus plot, genes and histone peaks colored in red had significant associations (FDR < 0.05) with the duplication.

Similarly, as observed with SV-eQTLs in the combined studies analysis (**Figure 3c**), the majority of SVs associated with one or more molecular traits were found near gene bodies. For instance, more than 87% of SVs associated with H3K9ac peaks (haSVs) had at least one breakpoint within 500 kb of the closest gene, while more than 93% of splicing associated SVs (sSV) were found within 50 kb of the respective gene bodies (**Supplementary Figure S9**). Additionally, the direction of effect for the associations (β) were usually distributed in both directions for SV-xQTLs, independently of SV class, reflecting possibly complex enhancing and repressing regulatory effects or loci with SVs in linkage disequilibrium (LD) with the true causal variants. However, when the SVs overlapped the phenotypes (e.g., exonic region or histone peak) the effects of deletions and MEIs were mostly negative, while duplications were mostly positive (**Supplementary Figure S10**), suggesting a likely causal role for SVs at these conditions.

By measuring associations for each SV class separately, we observed that specific classes were more likely to be associated than others in each phenotype. Deletions in particular showed enrichment of associations compared to all classes together, while insertions were depleted. *Alu* elements, despite being known to promote alternative splicing^32,33^, were enriched in eQTLs and pQTLs but not in the other two traits, while SVA elements were enriched in eQTL, pQTL and sQTLs (**Figure 4b**). SVAs are considerably less frequent than other transposable elements and their effects on splicing, expression, and protein could be due to SVAs acting as novel promoters^34^ or exon-trapping^35^.

SV-xQTLs were enriched in relevant functional annotations similarly across all molecular phenotypes (**Figure 4c**). SVs overlapping topological associated domains (TADs), exons, introns, and promoters were enriched for associations, while intergenic SVs were depleted. However, some specific phenotypes showed stronger enrichment than others. For instance, haSVs were strongly enriched in regulatory regions, such as promoters, enhancers, and CTCF sites relative to other phenotypes, highlighting the direct effect of SVs on the phenotype.

We identified 667 SV-gene pairs associated with at least two phenotypes with highly concordant effects. The correlation of effect sizes between eQTLs and pQTLs was 0.71 (Pearson correlation) and between pQTLs and haQTLs 0.77 (**Supplementary Figure S11**), while eQTLs and haQTLs showed slightly weaker correlation (Pearson correlation = 0.57) (**Figure 4d, Supplementary Figure S11**). In addition, 241 SVs were found affecting at least three phenotypes, and 28 SVs affecting all four measured phenotypes in several loci such as *HLA, GSTM, GSTT, RBM, BPHL, VARS2, CAB39L, RLBP1, GCSH, DECR2*, and *PHYHD1*. Moreover, more than 62% of SVs associated with proteins (pSVs) were also associated with differential RNA expression (**Figure 4e**). While the majority (87%) of the SV-pQTLs and SV-eQTLs were concordant (**Figure 4d**), few had discordant effects; for example, in the gene *UROS*, a 411 bp duplication located in the promoter region of the gene was associated with lower RNA expression, but higher protein expression, suggesting some complex regulatory mechanism (**Figure 4f**). Additionally, 25.5% and 23.7% of pSVs were also associated with histone markers and splicing, respectively, suggesting distinct mechanisms for gene regulation, while 7% of pSVs were associated with the four phenotypes, and 28% were found associated with proteins only (**Figure 4e**). By contrast, 50% and 47% of splicing and histone associated SVs were also SV-eQTLs, respectively (**Supplementary Figure S12**).

### Effects of rare SVs

In contrast to common variants which are widespread in a population and have been subjected to a long process of natural selection, rare variants are usually much more recent and their impact on phenotypes more deleterious^14,36^. Due to their low frequencies, the impact of rare variants is usually measured indirectly by looking for enrichments within outliers, instead of performing standard association tests^36,37^. To assess the impact of rare SVs in gene expression and gene regulation, we first mapped gene-sample expression outliers using *OUTRIDER*^*38*^, for RNA and protein levels measured for the ROS/MAP. Then, we assessed the enrichment of rare variant carriers nearby those genes.

We identified 1,551 and 1,747 gene-sample outlier pairs for RNA expression and protein levels, respectively (see Methods). A higher proportion of outliers was observed in proteins compared to RNA when considering samples and genes measured in common (112 samples and 7,546 genes) (**Figure 5a**). Additionally, only 43 (5%) gene-sample pairs were replicated between both phenotypes, reflecting the modest correlation (Spearman’s ρ = 0.38) observed between average RNA expression and protein levels (**Supplementary Figure S19**).

**Figure 5.**
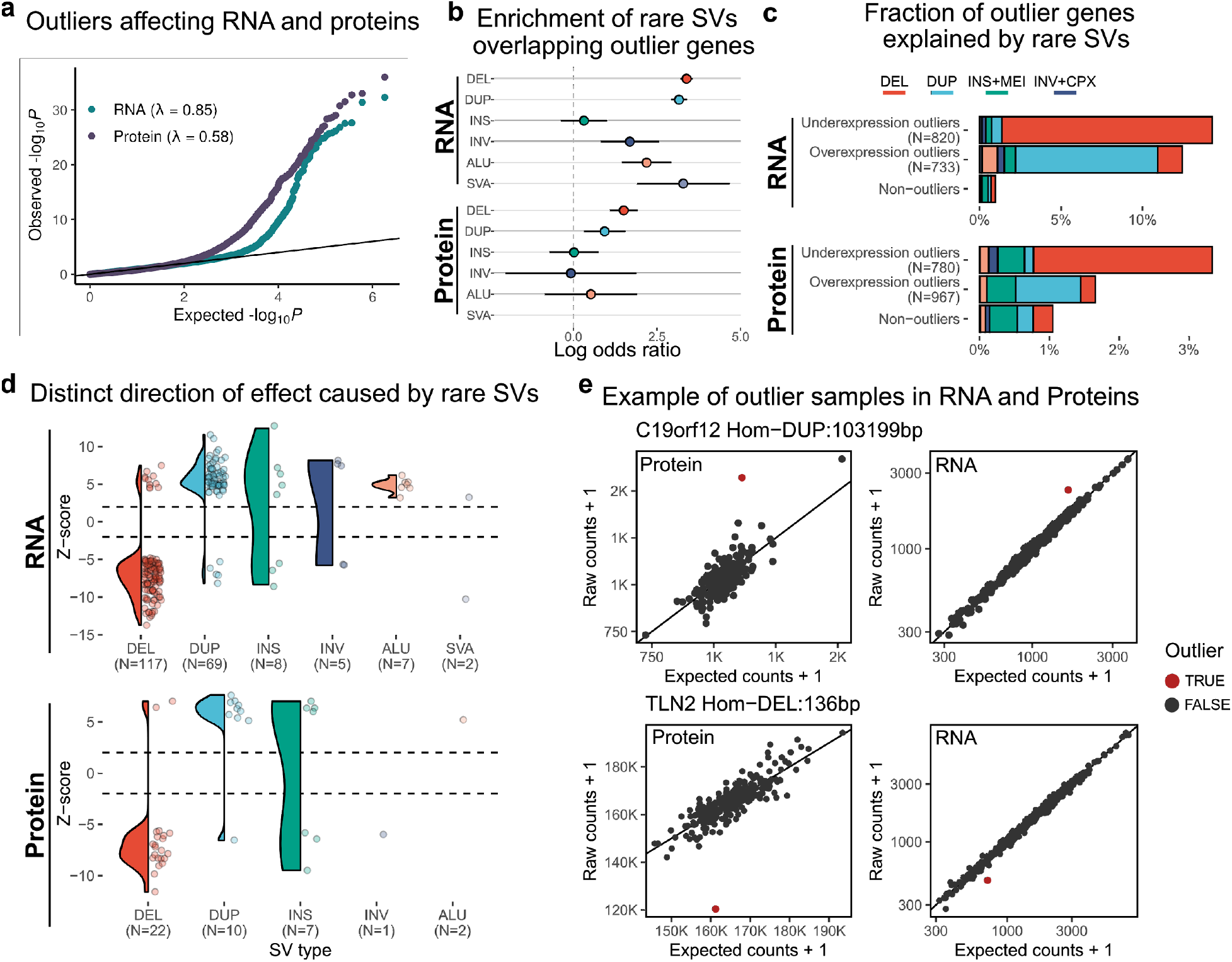
Impact of rare SVs on gene expression outliers. **a**, Quantile-quantile (QQ)-plot showing the observed distribution of *P*-values of outliers for RNA and protein and its deviation from the expected uniform distribution (showing only for gene-sample pairs measured in common). **b**, Enrichment of rare SVs overlapping outliers (any SV breakpoint within the gene body) stratified by SV type showed as a log odds ratio with 95% Wald confidence intervals. **c**, Fraction of overexpressed and underexpressed outlier genes that are potentially explained by each rare SV compared to non-outliers. **d**, Distribution of gene outlier z-scores that are overlapped by rare SVs. **e**, Examples of gene-sample pairs outlier with a rare SV overlapping their respective gene bodies. Showing on top an overexpression outlier for *C19orf12* caused by a 103 kb duplication and at bottom an underexpression outlier for the gene *TLN2* caused by a rare 136 bp deletion. Each dot represents a sample. Y-axis represents the raw counts + 1, while the x-axis represents the expected counts + 1, given by *OUTRIDER’* autoencoder model. Red dots represent an outlier sample.

Next, we measured the enrichment of rare SVs (MAF < 1%) overlapping gene bodies of outliers (for RNAs and proteins, separately). We found significant enrichment of SV classes in these conditions, especially deletions and duplications, with stronger enrichments in RNA compared to proteins (**Figure 5b**). This could be due to smaller sample sizes and the smaller number of genes tested. The direction of differential expression correlated with the expected effect caused by a given SV type (i.e., dosage alteration) (**Figure 5c**), but we still observed many cases in opposite directions suggesting more complex regulatory effects (**Figure 5d**). Six gene-sample outliers with overlapping rare SVs were found with effects on RNA and protein levels, including a homozygous rare 103 kb duplication causing overexpression of *C19orf12* and a homozygous 136 bp deletion causing underexpression of *TLN2* in the respective variant carriers (**Figure 5e**).

### Characterizing pathogenic SVs in neurodegenerative diseases

Since SVs are not usually included in GWAS, their association with neurodegenerative diseases and complex traits has been overlooked. Here we performed one of the first genome-wide SV associations with Alzheimer’s disease (AD) and progressive supranuclear palsy (PSP). By combining all SVs, we generated a combined call set with 29,177 SVs (22,007 with MAF > 1%) in 1,757 samples. In AD (539 cases, and 368 controls) no SVs were associated with the disease, however, some suggestive hits were observed (**Supplementary Figure S13**), including a 199 bp deletion downstream of the gene *CHRD* (nominal *P-*value 4.88 × 10^−5^), a locus previously reported associated in familial late-onset AD^39^. By contrast, for PSP (83 cases, 368 controls), identified four SVs after Bonferroni correction (**Figure 6a**). These variant alleles were highly correlated with each other and tagged known distinct haplotypes at the 17q21.31 locus defined by an almost 1 Mb inversion (**Figure 6b**). These haplotypes were previously reported to be associated with PSP and Parkinson’s disease, with the inverted haplotype being protective in both diseases (Odds ratio of 0.2 and 0.8 respectively)^40–43^. In addition, many of these SVs showed associations with changes in gene expression and other molecular phenotypes (**Figure 6c**). Of the associations replicated in at least one brain region across studies, we found higher expression of *DND1P1*, *KANSL1*, *ARL17A*, *LRRC37A* in the inverted haplotypes (**Figure 6c**) and differences in *MAPT* splicing junctions and several histone acetylation markers could be detected in ROS/MAP (**Figure 6c**). Recently a mechanism involving neuron-specific changes in chromatin accessibility and 3D interaction has been proposed^44^. However, additional studies are needed to demonstrate these effects on regulatory interactions.

**Figure 6.**
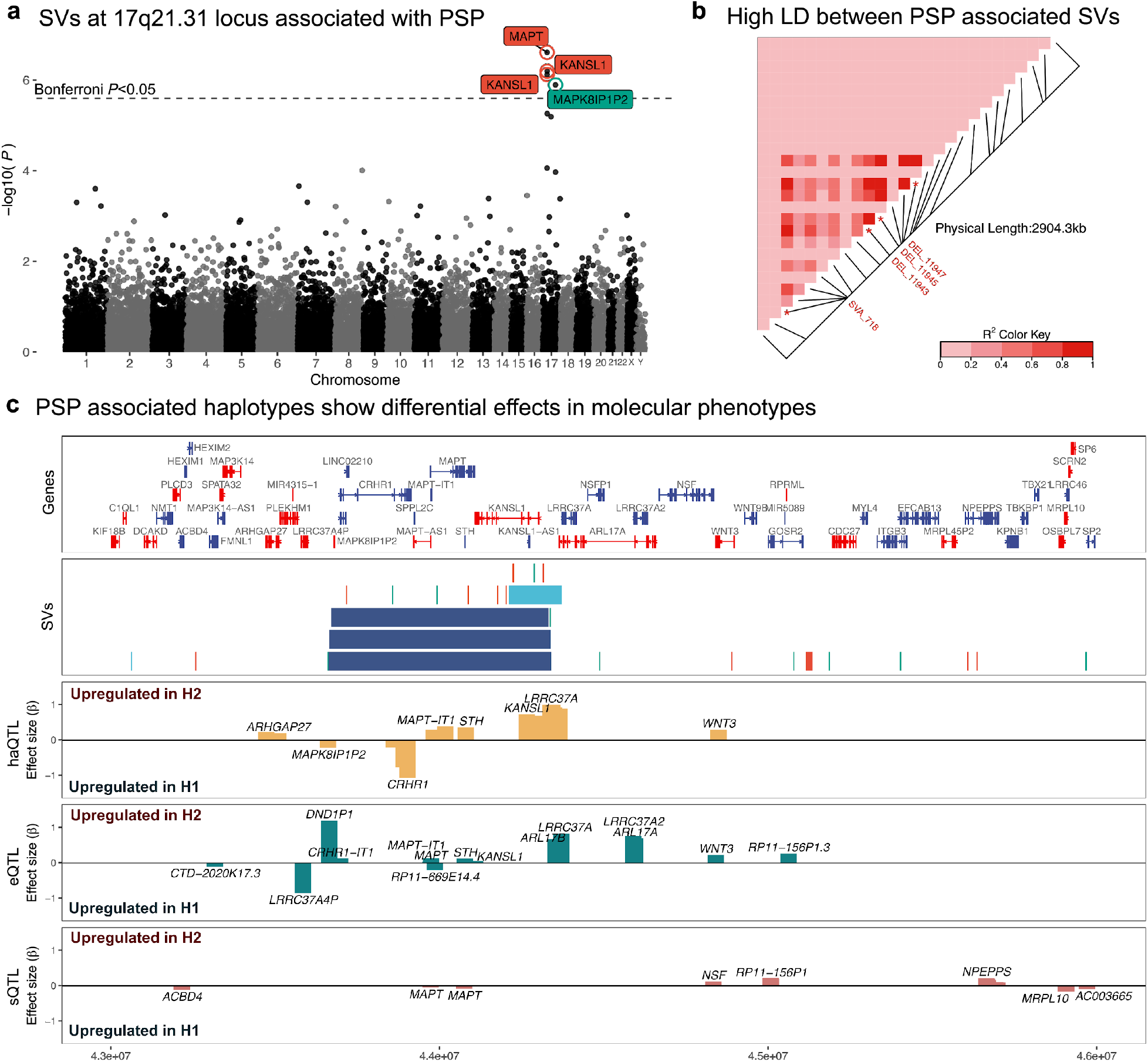
SVs associated with PSP and their effects on molecular phenotypes. **a**, Manhattan plot showing SVs associated with PSP cases (n=83) versus controls (n=368). Estimates were measured using Bayesian logistic regression (bayesglm) accounting for sex, study, and the first three ancestry PCs. Y-axis shows the -log_10_(*P*-value) of each SV association. X-axis represents SV sequential position by chromosome (not real scale). Labels with names of the nearest gene upstream of each SV breakpoint are shown for SVs with Bonferroni adjusted *P*-values lower than 5% (dashed line). Label colors represent different SV classes. **b**, Pairwise linkage disequilibrium (LD) matrix of SV genotypes identified between chr17:43M-46M (hg19) measured as R^2^ (LDheatmap R package). Labels are shown for the SVs significantly associated with PSP status (from letter a). **c**, Locus plot of 17q21.31 locus (chr17:43M-46M (hg19)). Genes bodies are shown at the top track, SVs with MAF ≥ 1% identified in ROS/MAP are shown at the second track (colors represent SV class), and effect sizes for H1-H2 inversion haplotypes (using the top PSP associated SVs - DEL_11943 - as a proxy) are shown in the remaining tracks. Effect sizes are shown only for significant associations (FDR < 0.05). Positive effect sizes indicate increased levels of each phenotype in individuals with H2 (inverted) haplotype.

## DISCUSSION

Detecting SVs accurately is a challenging task and limitations due to sample size and sequencing read length are the main challenges to the field^45^. Here we performed SV discovery integrating seven different algorithms designed to detect multiple classes of SVs using distinct sequencing features. Integration of tools was assisted using the Genome in a Bottle benchmark set^46^ and in addition, we generated long-read WGS for two brains and performed *in silico* confirmation of SVs predicted using short-read data. Our results showed improved sensitivity of SV detection compared to single algorithmic approaches as well as high orthogonal discovery confirmation on selected samples. Given the limitations of short-read data, SV discovery sensitivity is still underestimated for some SV classes, such as large insertions and complex configurations. However, we not only observed high reproducibility of SVs compared to independent large SV cohort studies and databases, but we also identified novel variants emphasizing the improvement of discovering SVs from novel samples and diverse populations. Importantly, we are making these resources publicly available.

Most studies on the impact of SVs have been restricted to the level of mRNA expression^1,14,30,47^. However, mRNA is not the only determinant of cellular functions^48^. Previous studies involving small variants found that QTL effects can be modulated at different levels of gene regulation^11–13^. Therefore, understanding the links between genetic and phenotypic variation is essential to understand how genetic variation, including SVs, impacts the regulation of gene expression. Here, we expanded such catalogs by measuring the impact of SVs through a regulatory cascade by integrating multiple ‘omics’ generated from histone acetylation markers up to gene expression and protein levels. We identified properties of SVs affecting different molecular phenotypes, identified regions and genes more susceptible to associations, and correlated their effects on phenotypes in terms of both common and rare SVs. Our SV-xQTLs results recapitulated similar trends from SNVs. For example, the majority of SVs associated with proteins were also SV-eQTLs, similar to what has been observed with SNV QTLs^11^. Although sQTLs and eQTLs tend to have independent lead variants in SNVs^12^, for SVs we observed that half of splicing SVs were also expression SVs, with a modest negative correlation between effect sizes. Additionally, many effects seemed to be specific to a phenotype with about 28% in SVs associated at protein level only which is 3-fold more than SNVs^11^. These data suggest that distinct mechanisms are involved in translating genotype to phenotype.

Interestingly, distinct SV classes seem to have different functional impacts on gene regulation. Transposable elements were shown to contribute to almost half of open chromatin regions^49^ and affect more than three fourths of promoter regions, with particular enrichment of short interspersed nuclear elements (SINE) (e.g., *Alu* elements)^50^. Here we found that *Alu* and SVA (composed of SINE-VNTR-Alu) elements are more likely to affect gene and protein expression compared to other SV classes. SVA elements in particular are more evolutionarily recent than other TEs and many are human-specific^34,51–53^. Their importance for gene expression were described both *in vitro* and *in vivo*^*54–57*^. Our results support an important role for SVA in gene regulation, with more than 2-fold greater chance of being associated with either gene expression, splicing, and protein levels (**Figure 4b**).

While most of the common SV-xQTL associations can be confounded by LD with actual causal SNVs^14^, rare SVs impacting expression outliers at RNA and protein levels can provide a better sense of SV causality^37^. Here we mapped genes with deviations from expected expression level distributions at mRNA and protein levels and then identified samples carrying rare SVs near those genes. We found more than 10% of mRNA outliers being overlapped by a rare SV, with clear causal resulting effect (e.g., deletions causing reduced expression while duplications causing increased expression). Interestingly, rare and common *Alu* elements seemed to have opposite effects on mRNA expression. Rare *Alu* insertions were found only in overexpression outliers (**Figure 5d**), while common *Alu* carriers were mostly associated with decreased expression (**Figure 3e**). Additionally, effects of rare SVs seem to be attenuated at protein levels, given a lower proportion of outliers explained by nearby SVs and an even lower proportion of effects shared between RNA and proteins, reflecting low correlation observed in the expression levels (**Supplementary Figure S19**).

In summary, our study expands the catalog of high-quality SVs by measuring their impact through a gene regulatory cascade. Our multiple ‘omics’ integration demonstrates an important role of SVs, showing that the effects of SVs differ in terms of SV classes, genomic locations, genes biotypes, and phenotypes affected. Our comprehensive SV-xQTL catalog allowed us to examine the contribution of *cis*-acting structural variants on number of molecular traits and provide a powerful resource for understanding mechanisms underlying neurological diseases.

## METHODS

### Study cohorts

In our analysis, we included samples from four cohorts (ROS, MAP, MSBB, and Mayo Clinic) from the Accelerating Medicines Partnership in Alzheimer’s Disease (AMP-AD) consortium. These aging cohorts provide an extensive collection of multi-omics data, that includes deep whole-genome sequencing (WGS) from 1,860 subjects, and allow us to identify SVs and characterize their functional impact. Each cohort is briefly described below.

#### ROS/MAP

The Religious Orders Study (ROS) and Rush Memory and Aging Project (MAP) are clinical-pathological cohort studies of aging and dementia based at the Rush Alzheimer’s Disease Center. ROS subjects live in communities distributed throughout the U.S., while MAP subjects live in communities in the Chicago metropolitan area. Both studies recruit older persons without known dementia who agree to annual clinical evaluation and organ donation at death. Both studies were approved by an Institutional Review Board of Rush University Medical Center. All participants signed an informed consent, Anatomic Gift Act, and repository consent to allow their data to be shared. Much of the ROS/MAP data are harmonized at the item level, including all omics generated from the same pipelines and QC’d together allowing efficient merging of datasets. ROS/MAP data can be requested at online (see URL at Data Availability section).

#### Mayo Clinic

Also referred as ‘Mayo RNAseq’, the Mayo Clinic cohort is an independent study from those described under the Mayo Clinic Alzheimer’s Disease Genetics Studies (MCADGS) and consists of 349 human subject DNA samples from Mayo Clinic in “Project_SCH_11923”.

#### MSBB

The Mount Sinai Brain Bank (MSBB) cohort consists of 349 samples assembled after applying stringent inclusion/exclusion criteria and represents the full spectrum of disease severity. Subject samples were collected and sequenced in Mount Sinai Project_SCH_11923. DNA was isolated from 50 mg of frozen, never-thawed grey matter dissected from the frontal cortex (BM10) or superior temporal gyrus (BM22). Specimens were homogenized in a 300 μl of elution buffer, and DNA was isolated using a Promega Maxwell 16 semi-automated system using the Promega Maxwell 16 Tissue DNA Purification Kit according to the manufacturer’s instructions. DNA quality and yield were assessed using an Agilent 4200 TapeStation.

### Whole-genome sequencing

Human postmortem tissues were acquired under proper Institutional Review Board (IRB) protocols at each respective institution. For all cohorts, whole-genome sequencing (WGS) libraries were prepared using the KAPA Hyper Library Preparation Kit in accordance with the manufacturer’s instructions. Briefly, 650ng of DNA was sheared using a Covaris LE220 sonicator (adaptive focused acoustics). DNA fragments underwent bead-based size selection and were subsequently end-repaired, adenylated, and ligated to Illumina sequencing adapters. Final libraries were evaluated using fluorescent-based assays including qPCR with the Universal KAPA Library Quantification Kit and Fragment Analyzer (Advanced Analytics) or BioAnalyzer (Agilent 2100). Libraries were sequenced on an Illumina HiSeq X sequencer (v2.5 chemistry) using 2 x 150 bp cycles. Whole Genome data were processed with an NYGC automated pipeline. Paired-end 150 bp reads were aligned to the GRCh37 human reference using the Burrows-Wheeler Aligner (*BWA-MEM* v0.7.8) and processed using the *GATK* best-practices workflow that includes marking of duplicate reads by the use of Picard tools v1.83, local realignment around indels, and base quality score recalibration (BQSR) via Genome Analysis Toolkit (*GATK* v3.4.0).

### SV discovery pipeline

SV discovery was performed on each cohort using a custom pipeline integrating seven SV discovery tools. This pipeline is composed of several independent modules designed to run in a sequential manner. When possible, within each module, jobs can be run in parallel to speed up the analysis of large numbers of samples. Each module is described below.

- **Module 01: Sample QC** This module is based on the first module from the *HOLMES* pipeline^58^ and is designed to collect QC metrics from BAM files. For each sample, we estimate library complexity (*Picard EstimateLibraryComplexity*), collect alignment metrics (Picard CollectWgsMetrics, *Samtools flagstat*, and *Bamtools stats*), collect insert size metrics (*Picard CollectMultipleMetrics*), conduct sex inference, and check for aneuploidy.
- **Module 02: SV discovery** Including a comprehensive set of tools is crucial to extract the maximum possible information from sequencing data. Seven tools were included into the pipeline for SV discovery: *Delly* v0.7.9^59^, *LUMPY* v0.2.13^60^, *Manta* v1.5.0^61^, *BreakDancer* v1.4.5^62^, *CNVnator* v0.3.3^63^, *BreakSeq* v2.2^64^, and *MELT* v2.1.5^65^. Each tool comprises at least one of the three main approaches to detect SVs (i.e., read pairs (RP), split-read (SR), and read depth (RD). *Manta, Delly, LUMPY*, and *MELT* are based on RP and SR information, while *BreakDancer* uses RP only. CNVnator is based on RD information, and BreakSeq2 uses a breakpoint library mapping approach. In our pipeline, *BreakDancer, CNVnator*, and BreakSeq are run through the *SVE* pipeline v0.1.0^66^, while for the other ones the standalone tool is used.
- **Module 03: Consensus call set per sample** In order to integrate the results of different tools, we first tested several merging strategies and combinations and evaluated the quality of each with the Genome in a Bottle HG002 SV call set v0.6^67^ (details in Methods - *Structural variation performance benchmarking*). Based on all strategies evaluated, we choose the following merging rules for each SV type.
  - Deletion = *Manta* + (*LUMPY* + *Delly* + *BreakSeq* + *BreakDancer* + *CNVnator* – at least 2 callers support) – *(75% recall and 92% precision)*
  - Insertions = *Manta* + *BreakSeq* – *(22% recall and 95% precision)*
  - Duplications = *Delly* + *LUMPY* + *Manta* + *BreakDancer* + *CNVnator* (at least 2 callers support)
  - Inversions = *Delly* + *LUMPY* + *Manta* + *BreakDancer* (at least 2 callers support)
  - Translocations = *Delly* + *LUMPY* + *Manta* + *BreakDancer* (at least 2 callers support)
  To merge each tool we use *SURVIVOR*^*68*^ requiring 1000 bp maximum distance between breakpoints to merge SVs of the same type and on the same strand.
- **Module 04: Merge samples** SVs defined in the tool consensus step for each sample are merged into a population call set using *SURVIVOR*^*68*^considering 1000 bp distance between breakpoints, and taking into account SV classes and strands.
- **Module 05: Genotyping SVs in each cohort** After defining a joint populational call set in the cohort. The genotyping is performed separately for each sample and SV class (deletions, duplications, and inversions) using *smoove* “genotype” function (https://github.com/brentp/smoove) (a wrap function for *SVTyper*^*69*^). This step forces genotyping on all SVs for each sample by looking for evidence support from its respective alignment “.bam” files. Later, each genotyped sample is merged again into a single populational call set using *smoove* “paste”. Mobile Element Insertions (MEI) are identified, merged, and genotyped separately using *MELT* functions, and its results are harmonized with insertions found by the other tools after merging samples. For harmonization, we used a 70% reciprocal overlap criteria (considering their breakpoint positions and lengths) to convert insertions into MEIs.
- **Module 06: Post-discovery QC** After the genotyping step, outlier samples are identified and removed if they have SV counts higher than three times the interquartile range (IQR) for each SV class with more than 50 SVs on the average per genome. Samples blacklisted in previous steps due to sex mismatches or with missing metadata are also removed (details in *Sample QC prior SV discovery* and *Sample QC after SV discovery* sections). Next, SVs with more than 10% of missing genotype information or no ALT variants genotyped are filtered out. And finally, deletions and duplications with low read depth support indicated using *duphold*^*70*^ are removed (details in Methods - *Variant QC*).
- **Module 07: SV annotation** The final SV call set is annotated using *AnnotSV* v2.2^71^, which compiles functionally, regulatory, and clinically relevant information. For downstream analysis, we select only the “full” annotation, that reports all elements overlapping a given SV. Additional annotations such as DNase hypersensitive sites, three-dimensional chromatin architecture and CTCF binding sites obtained from <u>RuderferLab/CNV_FunctionalAnnotation (github.com)</u>^47^ are also integrated (details in Methods - *Functional genomic annotation sources*).

### Sample QC prior to SV discovery

Sample quality check was performed during different phases of the analysis. Prior to the SV discovery phase, we assessed the consistency between the WGS raw files and the metadata, and several sequencing and alignment metrics. From a total of 1,904 WGS raw “.bam” files, we found 43 samples with missing metadata information or some kind of duplication. We also collected WGS metrics for each sample, including library complexity (*Picard EstimateLibraryComplexity*), alignment metrics (*Picard CollectWgsMetrics, Samtools flagstat*, and *Bamtools stats*), insert size metrics (*Picard CollectMultipleMetrics*), sex inference, and aneuploidy check. Most of these analyses were based on the first module from the *HOLMES* pipeline^58^. We defined seven features to be used as sample filtering criteria, as follows:

- Discordance between reported and inferred sex: remove samples where the reported sex differed from inferred based on the fraction of reads aligned to chromosomes X and Y.
- Pairwise alignment rate: keep only samples where the fraction of read pairs successfully aligned were > 0.95.
- Chimera rate: keep samples with average chimeric rate < 0.04.
- Adapter rate: keep samples with average adapter rate < 0.07.
- Read length: keep samples with mean read length > 125 bp.
- Coverage: keep samples with mean sequencing coverage > 20%.
- Autosomal ploidy spread: keep samples where the absolute difference between the highest and lowest ploidy estimates from any two autosomal chromosomes were < 1.

As these cohorts have been somewhat previously QCed, the majority of the samples showed no discrepancies using these metrics, with the exception of some sex mismatches (30 samples for ROS/MAP and 5 samples for MSBB), and some abnormal ploidy and chimeric rate (22 samples in ROS/MAP). These samples were flagged and then removed in the downstream analysis (**Supplementary Table S5**).

### Sample QC after SV discovery

After the SV discovery phase, we applied additional sample filtering steps to exclude outlier samples. For each sample, we counted the total number of non-reference autosomal biallelic SVs for each SV class (DEL, DUP, INV, INS). Samples were labeled as outliers and removed from the downstream analysis if they had an SV count more than three times the interquartile range (IQR) beyond the third quartile, or less than six times the IQR below the first quartile for each SV class. Samples were also removed if the genotype missingness was higher than 10% (**Supplementary Table S1, Supplementary Figure S15**).

### Variant QC

Following the SV genotyping step, SVs with more than 10% of missing genotype information or no ALT variants genotyped in all samples were removed. Additionally, unbalanced SVs (i.e., deletions and duplications) were filtered by the relative fold-change of depth, given by *duphold*^*70*^. Two measures are calculated based on the guanine-cytosine (GC) content:

- DupHold Bin Fold-Change (DHBFC): fold-change for the variant depth relative to bins in the genome with similar GC-content.
- DupHold Flank Fold-Change (DHFFC): fold-change for the variant depth relative to flanking regions.

According to duphold evaluations on the Genome in a Bottle benchmark, DHFFC works best for deletions and DHBFC works slightly better for duplications^70^. Therefore, we applied the recommended thresholds of DHFFC < 0.7 for deletions and DHBFC > 1.3 for duplications. We evaluated these metrics after merging samples and removed any SV in which less than 70% of samples failed the given thresholds.

### Functional genomic annotation sources

In addition to the default annotations provided in the *AnnotSV* tool, we also integrated brain relevant annotations following previous studies^47^. Using scripts from <u>RuderferLab/CNV_FunctionalAnnotation (github.com)</u>, we downloaded and annotated our SVs in each cohort. Briefly, gene structure information was obtained from Ensembl v75. Open chromatin regions measured by DNase hypersensitive sites were downloaded from Roadmap Epigenomics^72^. Three-dimensional chromatin architecture using TAD domains identified using Hi–C from the prefrontal cortex were obtained from PsychENCODE^73^. CTCF binding sites from ChIP-seq from brain-relevant cell types were downloaded from ENCODE^74^ and overlapping peaks from each tissue were merged into a single consensus region (n = 100,894). Additionally, we also integrated ROS/MAP H3K9ac peaks into the annotation of each SV.

### Population ancestry assignments

We estimated ancestries of all individuals with *somalier* (https://github.com/brentp/somalier), using the raw “.bam” files as input. In short, *somalier* extracts a list of known polymorphic sites by accessing the exact base information from alignment files and classifies each sample ancestry using a neural network trained from a set of labeled samples. Individuals from our cohorts were assigned to one of the five super population labels obtained from 1000 Genomes Project high coverage data (African [AFR], Ad Mixed American [AMR], East Asian [EAS], European [EUR], South Asian [SAS]) and were projected using principal component analysis (PCA).

### Reproducibility of SVs in other large cohort studies

SVs discovered in the AMP-AD cohorts were compared with other large cohort studies and datasets in order to identify novel variants. SV annotations were obtained from *AnnotSV* v2.1^71^ and included dbVar^26^, the National Human Genome Research Institute (NHGRI) Centers for Common Disease Genomics (CCDG)^10^, Database of Genomic Variants (DGV)^27^, Deciphering Developmental Disorders (DDD)^28^, GnomAD-SV^9^, and 1000 Genomes Project SVs^1^. SVs were considered replicated in other datasets if their coordinates had a reciprocal overlap of 0.7 irrespectively of the SV class.

### Structural variation performance benchmarking

In order to measure the performance of individual SV detection tools and merging strategies, we applied the SVs benchmarking dataset from GiaB v0.6 of Tier 1 (isolated, sequence-resolved SVs)^67^. This dataset contains SVs for the AshkenazimTrio son HG002 (NA24385) defined using several sequencing strategies and SV detection tools as well as a manual curation. For measuring the precision and recall for each method, we used the tool *truvari* (<u>spiralgenetics/truvari: Structural variant toolkit for VCFs (github.com)</u>).

Results for each tool were pre-filtered before comparisons by keeping only SVs from canonical chromosomes (1-22 plus X and Y), removing SVs <= 50 bp, and keeping only PASS filters only (**Supplementary Table S2**). Next, precision and recall for deletion and insertion discovery were evaluated for each tool. This initial comparison showed how discordant and inefficient the SV discovery using short-reads can be (**Supplementary Figure S14a**).

To improve the results of individual tools, we tested different combinations for each SV type. We applied 5 merging strategies: 1) union of all tools; 2) intersection of all tools; 3) merging calls detected by at least ‘n’ tools; 4) union of a combination of specific tools plus calls detected by at least ‘n’ other tools; and 5) intersection of tools plus calls detected by at least ‘n’ other tools. The best merging strategy was then selected after measuring all possible combinations for each strategy separately for DEL and INS and selecting the best results based on the F1-score measure. Thus, for deletions, the best performance was found by adopting all deletions from *Manta* plus all other deletions found by at least two other tools (**Supplementary Table S3**). While for insertions, the best F1-score was obtained using a union of calls from *Manta* and *BreakSeq* only (**Supplementary Table S3**). Using these combinations we were able to improve the overall recall from 41% (*Manta*-only) to 45% while keeping the precision at 88% (**Supplementary Figure S14a**). Since there are no benchmarking calls established yet on GiAB for other SV types (e.g., inversions and translocations), we took a more conservative approach and decided to keep only calls that were found by at least two different tools in the final merged set. After applying this strategy to all individuals in the AMP-AD cohorts, we measured their performance against the HG002 sample (**Supplementary Figure S14b**). This analysis showed similar performances across all individuals, with expected reduced precision since variants between different individuals were being compared. The complete report of all analysis tests is available at <u>RajLabMSSM/StructuralVariation (github.com)</u>.

### Allele frequency comparison with 1000 Genomes Project and gnomAD-SV

Correlation of allele frequencies (AFs) between SVs discovered in gnomAD-SV and 1000 Genomes Project phase III were compared to ROS/MAP AFs. Only European (EUR) AFs from gnomAD and 1000 Genomes were used for comparison. SVs in common were first identified using *bedtools* “intersect” requiring at least 50% reciprocal overlap with no requirement of matching SV classes. Then, coefficients of determination (R^2^) were assessed with a linear regression between log_10_ of AFs for SVs with AF > 1% in both studies being compared (**Supplementary Figure S4**).

### Hardy-Weinberg equilibrium comparison

SV genotype distributions were evaluated under the null expectations set by the Hardy-Weinberg equilibrium (HWE; 1 = p^2^ + 2pq + q^2^). Using tabulated genotype distributions per cohort as input, we measured deviations from HWE using a chi-square goodness-of-fit test with one degree of freedom and their *P*-values using the “HardyWeinberg” package in R^75^. SVs were considered in violation of HWE if its *P*-value was significant after Bonferroni correction for the number of SVs tested per population (**Supplementary Figure S5**).

### SV long-read validation

Selection of samples for long-read validation was performed using SVCollector^76^. Samples were initially ranked using both, the “topN” (i.e., sample with the largest number of SVs, irrespectively of other samples) and the “greedy” (i.e., samples that collectively contain the largest number of distinct SVs) approaches. Two samples from ROS/MAP cohorts were selected considering the best overall ranking in both approaches and the existence of additional omics data for those samples (e.g., RNA, protein, and histone markers) (**Supplementary Table S4**).

DNA samples extracted from DLPFC tissues were then used for continuous long-read (CLR) sequencing using PacBio Sequel II platform. Both samples were multiplexed sequenced in a single SMRT Cell 8M Tray, resulting in an average 10x coverage per sample and average 14 kb read length (**Supplementary Table S5**). Raw PacBio BAM files were then aligned to the GRCh37 reference genome using *minimap2*^*77*^ and were used to validate SVs found using the orthogonal short-read data using *VaPoR*, a software that performs comparative local realignments of long-reads to a synthetically modified reference sequence^78^.

Therefore, SVs identified in the main SV discovery step with short-reads and positively genotyped in each sample were selected and filtered to maximize *VaPoR* sensitivity. We restricted the analysis for SVs with no overlapping breakpoints to simple repeats, segmental duplications, centromeres, regions subject to somatic V(D)J recombinations, and regions with low mappability in the PacBio data (<10x coverage). SV classes were evaluated separately by deletions, duplications, and insertions. For inversions, since our calls were not completely resolved and could represent also other sorts of complex conformations, we measured their support either as simple inversions (INV) or any combination of deletions, duplications, and inversions (e.g DEL_INV, DUP_INV, DEL_DUP_INV). SVs with a proportion of reads supporting the predicted structure versus all reads assessed higher than zero (i.e., VaPoR_gs > 0) or SVs with genotype proposed by *VaPoR* other than homozygous to the reference (i.e., 0/0) were considered supported in the long-read data. Supporting rates for each sample were then measured as the number of supported SVs divided by the total number of tested SVs (**Figure 2d**).

### RNA-seq processing and SV-eQTL mapping

Given that originally each cohort had different RNA-seq processing pipelines, we took advantage of the RNA-seq Harmonization Study (rnaSeqReprocessing) data (<u>Synapse:syn9702085</u>), which reprocessed all the data in a harmonized workflow. The reprocessing was done using a consensus set of tools with only library type-specific parameters varying between pipelines. Briefly, in this study, FASTQ files were obtained from aligned BAM files converted into raw FASTQs using *Picard SamToFastq*. Then, sequences were aligned to the GENCODE25 (GRCh38) reference genome using STAR and gene counting was computed using STAR option *quantMode* set as *GeneCounts*. Later, in order to match the WGS reference (GRCh37) used for SV discovery in the present study, gene coordinates were lifted-over using the Gencode annotations (“gencode.v24lift37.annotation.gtf”). For SV-eQTL analysis, we used previously processed residuals values obtained after adjusting for clinical, technical, and hidden (via SVA^79^) confounders following conditional quantile normalization (CQN) normalization to account for variations in gene length and GC content and outlier detection and removal. We mapped SV-eQTL to scan for significant associations between common structural variants and gene expression. We tested SVs with MAF ≥ 0.01 using a modified version from *FastQTL*^*14,80*^ to address the span of breakpoints within a 1 Mb window from each gene TSS. A permutation test was applied to select the lead SV per gene and *P*-values were adjusted for multiple testing using Benjamini-Hochberg (FDR). Associations were performed separately for each SV class, meaning that multiple lead-SVs (from different classes) could be associated with each phenotype.

### SV-haQTL mapping

ChIP-seq experiments and data processing for H3K9ac acetylation markers were previously performed on 712 samples (699 after QC) Epigenetics (ChIP Seq) - syn4896408 (synapse.org)^81^. Briefly, short single-end reads were aligned against the GRCh37 using the BWA. Peaks were detected for each sample individually by MACS2 using the broad peak option, a stringent q-value cutoff of 0.001, and pooled genomic DNA as a negative control library. Subsequently, H3K9ac domains were defined as genomic regions that were detected as a peak in at least 100 (15%) of our 669 samples. Regions neighboring within 100 bp were merged and very small regions of less than 100 bp were removed, resulting in 26,384 H3K9ac domains. H3K9ac levels were quantified for each sample and domain by counting the number of reads in the domains after extending reads towards the 3’-end by the estimated DNA fragment length. For SV-haQTL analysis, we used residualized values obtained from 571 samples with WGS after regressing out “Sex”, “gel_batch”, “AgeAtDeath” and the first 3 principal components of the genotype matrix to account for the effect of ancestry plus the first 10 principal components of the phenotype matrix to account for the effect of known and hidden factors (**Supplementary Figure S16**). We tested SVs with MAF ≥ 0.01 and within 1 Mb of each peak. A permutation test was applied to select the lead SV per peak. Finally, *P*-values were adjusted for multiple testing using Benjamini-Hochberg (FDR). Associations were performed separately for each SV class, meaning that multiple lead-SVs (from different classes) could be associated with each phenotype.

### SV-pQTL mapping

Tandem Mass Tag (TMT) isobaric labeling data were previously generated for 292 individuals^82,83^. For SV-pQTL analysis, we used residualized values for 7,960 protein obtained from 272 samples with WGS after regressing “PMI”, “Sex”, “AgeAtDeath”, three first ancestry PCs, and the first 10 principal components of the phenotype matrix (**Supplementary Figure S17**). We tested SVs with MAF ≥ 0.01 and within 1 Mb of each protein. A permutation test was applied to select the lead SV per protein. Finally, *P-*values were adjusted for multiple testing using Benjamini-Hochberg (FDR). Associations were performed separately for each SV class, meaning that multiple lead-SVs (from different classes) could be associated with each phenotype.

### SV-sQTL mapping

Splicing junction proportions, measured as percent spliced in (PSI), were measured previously^84^. Briefly, 110,092 junctions in 549 samples from ROS/MAP cohorts were obtained using *Leafcutter*^*85*^. Values were then standardized across individuals and quantile normalized across introns. For SV-sQTL analysis, we regress out the effects of known and hidden factors as performed in the sQTL manuscript^84^. We accounted for the effect of ancestry given by the first three principal components of the genotype matrix plus the first 15 principal components of the phenotype matrix (PSI) (**Supplementary Figure S18**). A total of 505 samples with WGS data were used in the association analysis using a modified version from *FastQTL*^*14,80*^ to address when the span or breakpoint of deletions, duplications, inversions, or insertions felt within the *cis* window a gene TSS. Genotyping information of SVs with MAF ≥ 0.01 and within 100 kb of each intron junction were tested, and a permutation test was applied to select the top SV per junction. Finally, *P*-values were adjusted for multiple testing using Benjamini-Hochberg (FDR). Associations were performed separately for each SV class, meaning that multiple lead-SVs (from different classes) could be associated with each phenotype.

### Expression outliers assessment

To identify expression outliers, either at RNA and protein levels, we used the *OUTRIDER* R package^38^. Briefly, data normalization was first performed using its inbuilt autoencoder method to control for variation linked to unknown factors. Then outlier detection was performed assuming a significant deviation of gene expression distributions from a negative binomial distribution. For the RNA, read counts for 15,004 genes expressed in 456 samples were used as input. While for proteins, we used the rounded batch adjusted abundances for 8,179 proteins and 272 samples. Samples with missing protein abundance values were imputed as the mean values of each protein. The significance threshold for outlier detection was set at FDR adjusted *P*-values of 0.05 and 0.001 for RNA and protein respectively and absolute z-scores higher than 2 for both data. A total of 1,551 gene-sample pairs outliers were identified in RNA, and 1,747 in proteins at the given thresholds.

### Enrichment analysis

All enrichments of SV features were accessed via logistic regression and adjusted by SV size. Briefly, data were converted to a binary matrix with lines representing each SV and columns representing related features (e.g., association to specific phenotype, overlapping genomic annotation, etc). Logistic regression was performed fitting a generalized linear model (*glm* R function) and log odds ratio estimates and *P*-values were extracted from each comparison.

### Disease status associations

SV calls from ROS/MAP, Mayo Clinic and MSBB were merged into a combined call set using *SURVIVOR*^*68*^ while requiring 1000 bp maximum distance between breakpoints to merge SVs of the same type. A total of 22,007 SVs identified and all three study groups and with MAF ≥ 0.01 were selected for the association test. Alzheimer’s disease status was harmonized across cohorts as previously described^86^. Briefly, for the ROSMAP study, late-onset AD (LOAD) cases was defined as individuals with a Braak neurofibrillary tangle (NFT) score ≥ 4, CERAD score ≤ 2, and a cognitive diagnosis of probable AD with no other causes, while individuals with Braak less ≤ 3, CERAD score ≥ 3, and cognitive diagnosis of “no cognitive impairment” were considered as controls. For MSBB, individuals CDR score ≥ 1, Braak score ≥ 4, and CERAD neuritic and cortical plaque score ≥ 2 were considered LOAD cases, while CDR scores ≤ 0.5, Braak ≤ 3, and CERAD ≤ 1 were considered controls (note that CERAD definitions differ between ROSMAP and MSBB studies). For the Mayo Clinic study, cases were defined based on neuropathology, with LOAD cases being individuals with Braak score ≥ 4 and CERAD neuritic and cortical plaque score > 1 while controls were defined as Braak ≤ 3, and CERAD < 2. A logistic regression was fitted using 539 AD cases and 368 controls and adjusting for sex, study, and the first three ancestry principal components. For PSP associations, Mayo Clinic study had 83 cases^87^ with pathological diagnosis at autopsy were compared against the same 368 controls using the same model.

## Data Availability

Code Availability: All code used in this study has been provided in a single repository on GitHub (https://github.com/RajLabMSSM/AMP_AD_StructuralVariation).
Data Availability: Data supporting the findings of this study are available from the AMP-AD Knowledge Portal (https://doi.org/10.7303/syn2580853). Specifically, whole-genome sequence data used for SV discovery are available for each cohort respectively: ROS/MAP (syn10901595); MSBB (syn10901600) and Mayo Clinic (syn10901601). ROS/MAP H3K9ac ChiP-seq data are available at syn4896408 and TMT proteomics data are available at syn17015098. RNA-seq reprocessed data from all cohorts were obtained from the RNAseq harmonization study (syn9702085). Splicing junction proportions were obtained from Raj et al. and a respective sQTL visualization (Shiny App) browser is available at https://rajlab.shinyapps.io/sQTLviz_ROSMAP/. ROSMAP data can also be requested at https://www.radc.rush.edu. All SV site-frequency data from 1,706 donors discovered separately in each cohort, complete nominal and permuted SV-xQTL summary statistics, and disease status association summary statistics are available at on GitHub (https://github.com/RajLabMSSM/AMP_AD_StructuralVariation).

https://doi.org/10.7303/syn2580853

## METHODS & SUPPLEMENTAL INFORMATION

Detailed methods and supplemental information for this manuscript has been provided online.

## ACKNOWLEDGMENTS

We thank the participants of AMP-AD cohorts for their essential contributions and gift to these projects. ROSMAP study data were provided by the Rush Alzheimer’s Disease Center, Rush University Medical Center, Chicago. Data collection was supported through funding by National Institute on Aging (NIA) grants P30AG10161, R01AG15819, R01AG17917, R01AG30146, R01AG36836, U01AG32984, U01AG46152, U01AG61356, and the Illinois Department of Public Health. Mayo RNA-seq study data were provided by the following sources: the Mayo Clinic Alzheimer’s Disease Genetic Studies, led by Dr. Nilufer Ertekin-Taner and Dr. Steven G. Younkin, Mayo Clinic, Jacksonville, Florida, using samples from the Mayo Clinic Study of Aging, the Mayo Clinic Alzheimer’s Disease Research Center, and the Mayo Clinic Brain Bank. Data collection was supported through funding by NIA grants P50 AG016574, R01 AG032990, U01 AG046139, R01 AG018023, U01 AG006576, U01 AG006786, R01 AG025711, R01 AG017216, and R01 AG003949; National Institute of Neurological Disorders and Stroke (NINDS) grant R01 NS080820; the CurePSP Foundation; and support from Mayo Foundation. Study data include samples collected through the Sun Health Research Institute Brain and Body Donation Program of Sun City, Arizona. The Brain and Body Donation Program is supported by the National Institute of Neurological Disorders and Stroke (U24 NS072026, National Brain and Tissue Resource for Parkinson’s Disease and Related Disorders), the NIA (P30 AG19610, Arizona Alzheimer’s Disease Core Center), the Arizona Department of Health Services (contract 211002, Arizona Alzheimer’s Research Center), the Arizona Biomedical Research Commission (contracts 4001, 0011, 05-901, and 1001 to the Arizona Parkinson’s Disease Consortium), and the Michael J. Fox Foundation for Parkinson’s Research. Mount Sinai Brain Bank (MSBB) data were generated from post-mortem brain tissue collected through the Mount Sinai VA Medical Center Brain Bank and were provided by Dr. Eric Schadt of the Mount Sinai School of Medicine through funding from NIA grant U01AG046170. The authors thank Dr. Bin Zhang and Dr. Erming Wang for assistance with data sharing, and members of the Raj and Crary labs for their feedback on the manuscript. We thank Jack Humphrey for his insightful comments and suggestions during this work. This work was supported by grants from the US National Institutes of Health (NIH NIA U01 AG06888001, NIA R01-AG054005, NIA R56-AG055824, and NIA R01-AG054008). This work was supported in part through the computational and data resources and staff expertise provided by Scientific Computing at the Icahn School of Medicine at Mount Sinai. We thank the Mount Sinai Technology Development core for help and support with performing long-read sequencing. The research reported in this paper was supported by the Office of Research Infrastructure of the National Institutes of Health under award number S10OD026880.

## AUTHOR CONTRIBUTIONS

Conceptualization: TR, RAV; Methodology: TR, RAV; Software: RAV; Formal analysis: RAV, KPL; Resources and data curation: DAB, TR, JFC; Writing - original draft: TR, RAV; Writing - review & editing: TR, DAB, JFC, RAV, KPL; Supervision, project administration, and funding acquisition: TR. All authors read and approved the manuscript.

## RESOURCE AVAILABILITY

### Code Availability

All code used in this study has been provided in a single repository on GitHub (https://github.com/RajLabMSSM/AMP_AD_StructuralVariation).

### Data Availability

Data supporting the findings of this study are available from the AMP-AD Knowledge Portal (https://doi.org/10.7303/syn2580853). Specifically, whole-genome sequence data used for SV discovery are available for each cohort respectively: ROS/MAP^88^ (syn10901595); MSBB^89^ (syn10901600) and Mayo Clinic^90^ (syn10901601). ROS/MAP H3K9ac ChiP-seq data are available at syn4896408 and TMT proteomics data are available at syn17015098. RNA-seq reprocessed data from all cohorts were obtained from the RNAseq harmonization study^86^ (syn9702085). Splicing junction proportions were obtained from Raj et al.^84^ and a respective sQTL visualization (Shiny App) browser is available at https://rajlab.shinyapps.io/sQTLviz_ROSMAP/. ROSMAP data can also be requested at https://www.radc.rush.edu. All SV site-frequency data from 1,706 donors discovered separately in each cohort, complete nominal and permuted SV-xQTL summary statistics, and disease status association summary statistics are available at on GitHub (https://github.com/RajLabMSSM/AMP_AD_StructuralVariation).

