## Supplementary Material for "The impact of genomic structural variation on the transcriptome, chromatin, and proteome in the human brain"

### LIST OF FIGURES

Supplementary Figure S1: SV discovery pipeline 4
Supplementary Figure S2: Sample sequencing metrics quality control 5
Supplementary Figure S3: Study group genetic ancestry 6
Supplementary Figure S4: Allele frequency comparisons with other studies 7
Supplementary Figure S5: Hardy-Weinberg Equilibria of SVs across groups 8
Supplementary Figure S6: Functional context and evolutionary constraints 9
Supplementary Figure S7: Correlation of direction of effects of SV-eQTLs across brain regions 10
Supplementary Figure S8: SV-xQTL top hits 11
Supplementary Figure S9: SV distance to the gene body (SV-xQTL) 12
Supplementary Figure S10: SV-xQTL effect sizes 13
Supplementary Figure S11: Correlation between SV-xQTL effects 14
Supplementary Figure S12: SVs affecting multiple phenotypes in the regulatory cascade 15
Supplementary Figure S13: Alzheimer's disease SV associations 16
Supplementary Figure S14: Quality assessment of variant calling 17
Supplementary Figure S15: Samples removed 18
Supplementary Figure S16: ChIP-seq H3K9ac QC 19
Supplementary Figure S17: TMT proteomics QC 20
Supplementary Figure S18: Splicing QC 21
Supplementary Figure S19: Correlation between RNA and protein expression 22

**LIST OF TABLES**

Supplementary Table S1 - Sample quality control 23
Supplementary Table S2 - Results for SV discovery in HG002 using different tools 24
Supplementary Table S3 - Benchmarking results 25
Supplementary Table S4 - Samples selected for SV validation 26
Supplementary Table S5 - Long read sequencing metrics 27

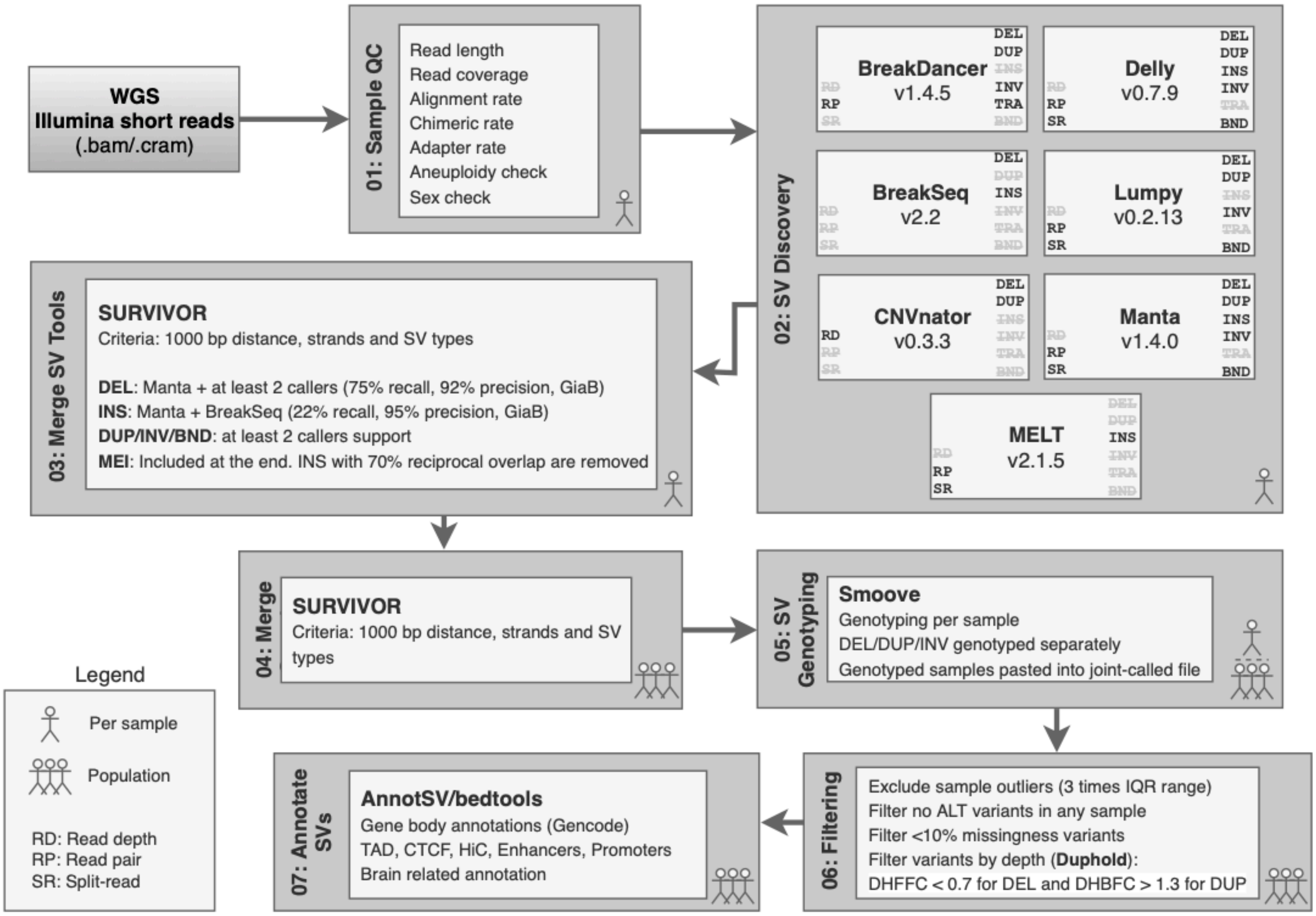


**Supplementary Figure S1: SV discovery pipeline.** The pipeline for SV discovery is composed of several independent modules designed to run in a sequential manner. Module 01 collects a series of sequencing quality metrics that are used to control low-quality samples in downstream analysis. Module 02 comprises seven different SV discovery tools that run in parallel for each sample. These tools cover different complementary methods that are further merged in the module 03 using *SURVIVOR*. In the modules, 04 and 05 SVs are jointly analyzed and genotyped using *smoove*. Module 06 performs sample outlier filtering and variant filtering according to several criteria thresholds. Finally, in module 07, the final SV call set is annotated using *AnnotSV* and *bedtools*.

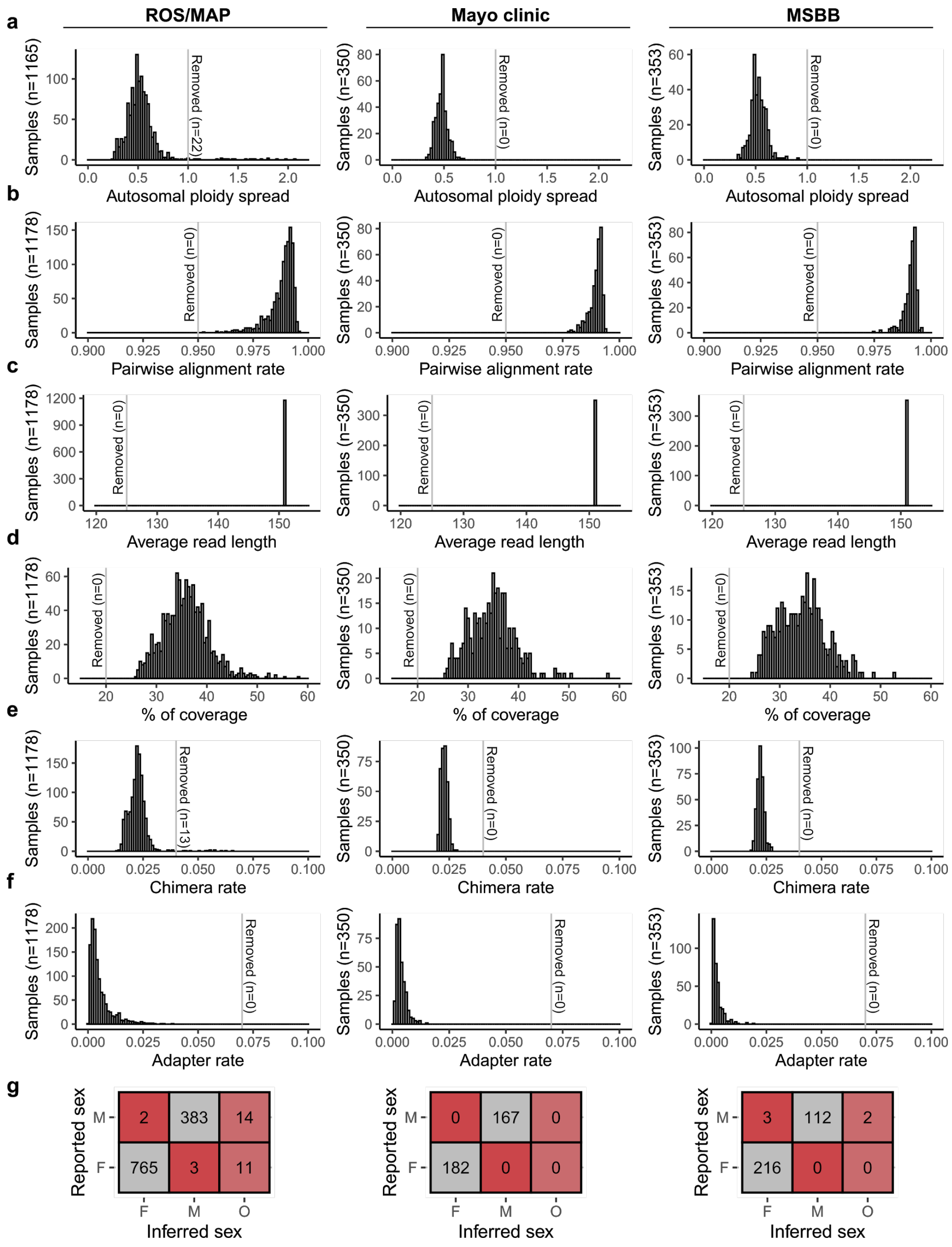


**Supplementary Figure S2: Sample sequencing metrics quality control.** Sample quality control performed prior SV discovery step. A series of sequencing quality metrics were collected from WGS ".bam" files. **a**, autosomal ploidy spread (keep samples where the absolute difference between the highest and lowest ploidy estimates from any two autosomal chromosomes were < 1). **b**, pairwise alignment rate (keep only samples where the fraction of read pairs successfully aligned were > 0.95). **c**, average read length (keep samples with mean read length > 125 bp). d, percentage of coverage (keep samples with mean sequencing coverage > 20%). **e**, chimera rate (keep samples with average chimeric rate < 0.04). **f**, adapter rate (keep samples with average chimeric rate < 0.07). **g**, sex mismatches (removal of samples with discordance between reported and inferred sexes).

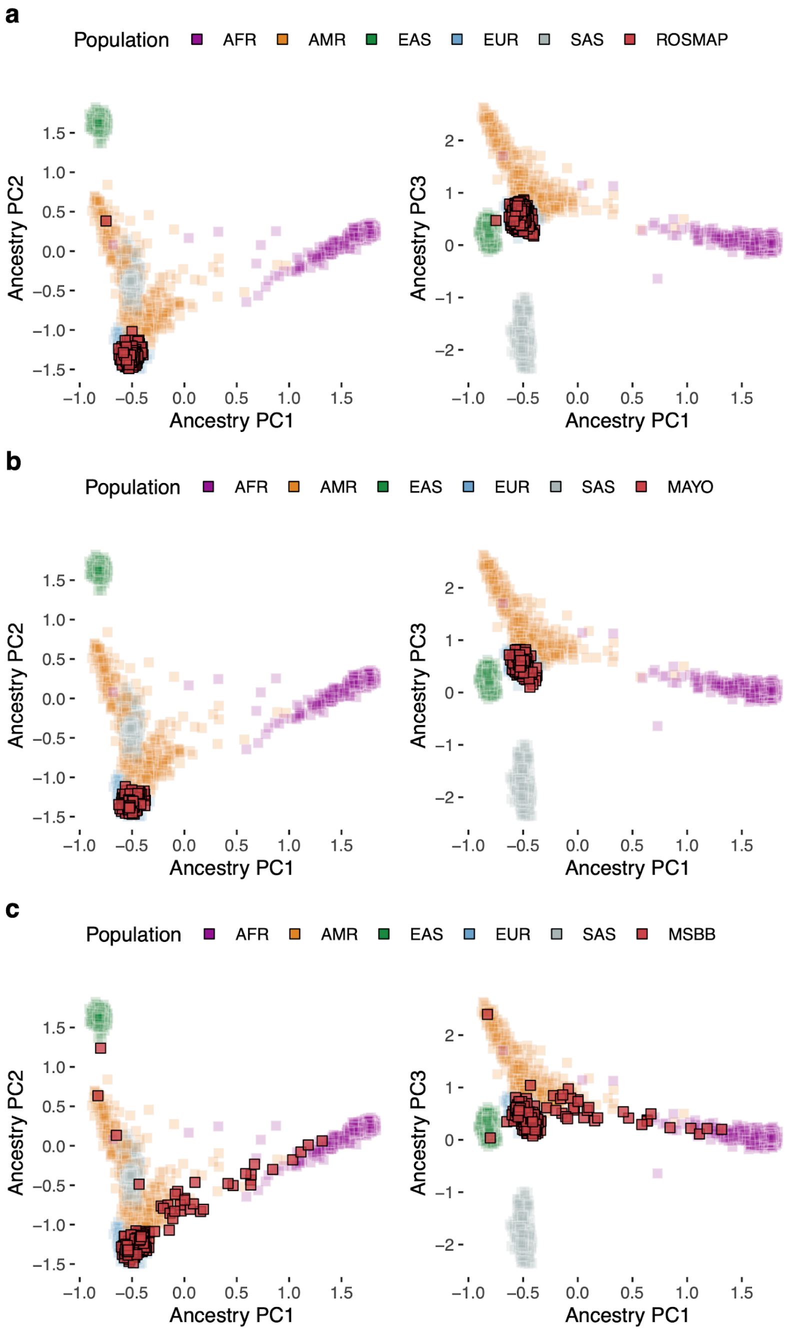


**Supplementary Figure S3: Study group genetic ancestry.** Ancestry prediction projected as principal component analysis for all individuals from: **a**, ROS/MAP; **b**, Mayo Clinic; and **c**, Mount Sinai Brain Bank (MSBB). Each group is plotted on top of 2,504 individuals from 1000 Genomes Project labeled in five super populations: African [AFR], Ad Mixed American [AMR], East Asian [EAS], European [EUR], South Asian [SAS].

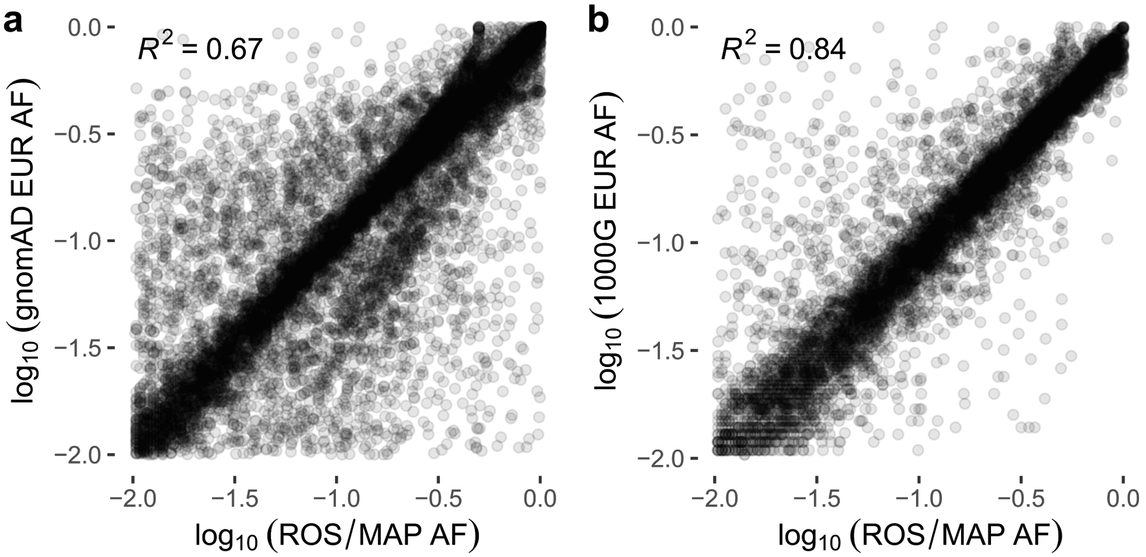


**Supplementary Figure S4: Allele frequency comparisons with other studies.** Correlation of allele frequencies (AFs) between SVs discovered in gnomAD-SV and 1000 Genomes Project phase III compared to ROS/MAP. **a**, 11,327 SVs with AF > 1% in both ROS/MAP (x-axis) and gnomAD (y-axis). **b**, 7,332 SVs with AF > 1% in both ROS/MAP (x-axis) and 1000 Genomes Project phase III (y-axis). Coefficients of determination ($R^2$) were assessed with a simple linear regression between $log_{10}$ of AFs. For comparison, only European (EUR) AFs were used from gnomAD and 1000 Genomes.

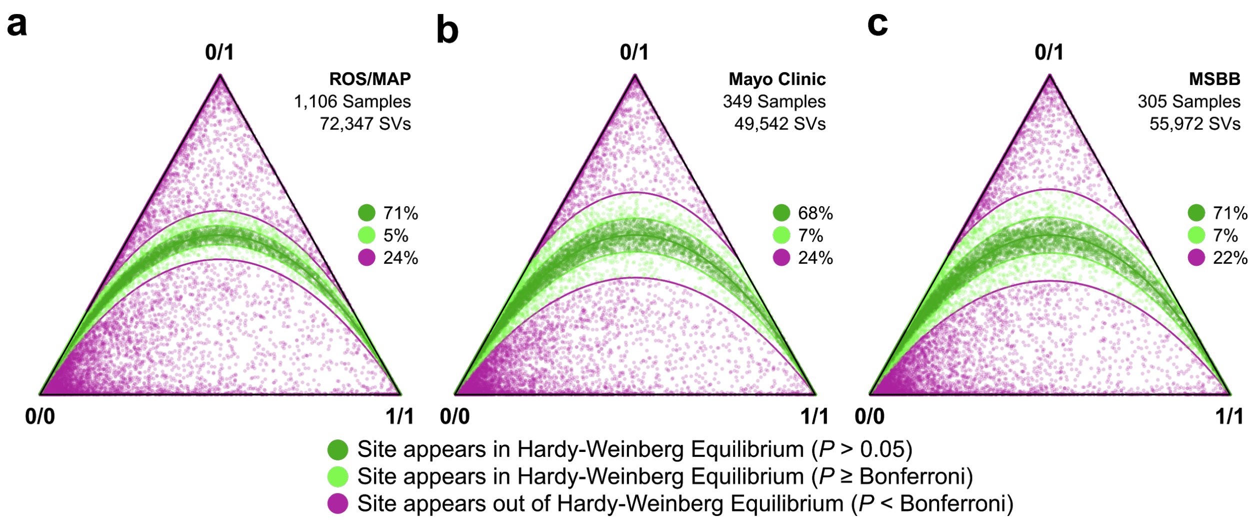


**Supplementary Figure S5: Hardy-Weinberg Equilibria of SVs across groups.** Hardy-Weinberg Equilibrium (HWE) statistics for each group: **a**, ROS/MAP; **b**, Mayo Clinic; **c**, MSBB. Each point is a single SV projected onto HWE ternary axes corresponding to its ratio of homozygous reference (0/0), heterozygous (0/1), and homozygous alternate (1/1) genotypes across all samples in the indicated group. The distance of a point to a vertex indicates the fraction of samples with that genotype. Deviation from HWE was assessed using a chi-square goodness-of-fit test with one degree of freedom, and points are colored based on their *P*-value. Green points are SVs within bounds defined for HWE based on the number of sites documented in each population, and purple points are SVs outside of these *P*-value bounds. The proportion of SVs corresponding to each *P*-value cutoff is provided at the right of each panel. Plots were generated using the "HardyWeinberg" package in R (Graffelman et al. 2015) and gnomAD-SV scripts (Collins et al. 2020)

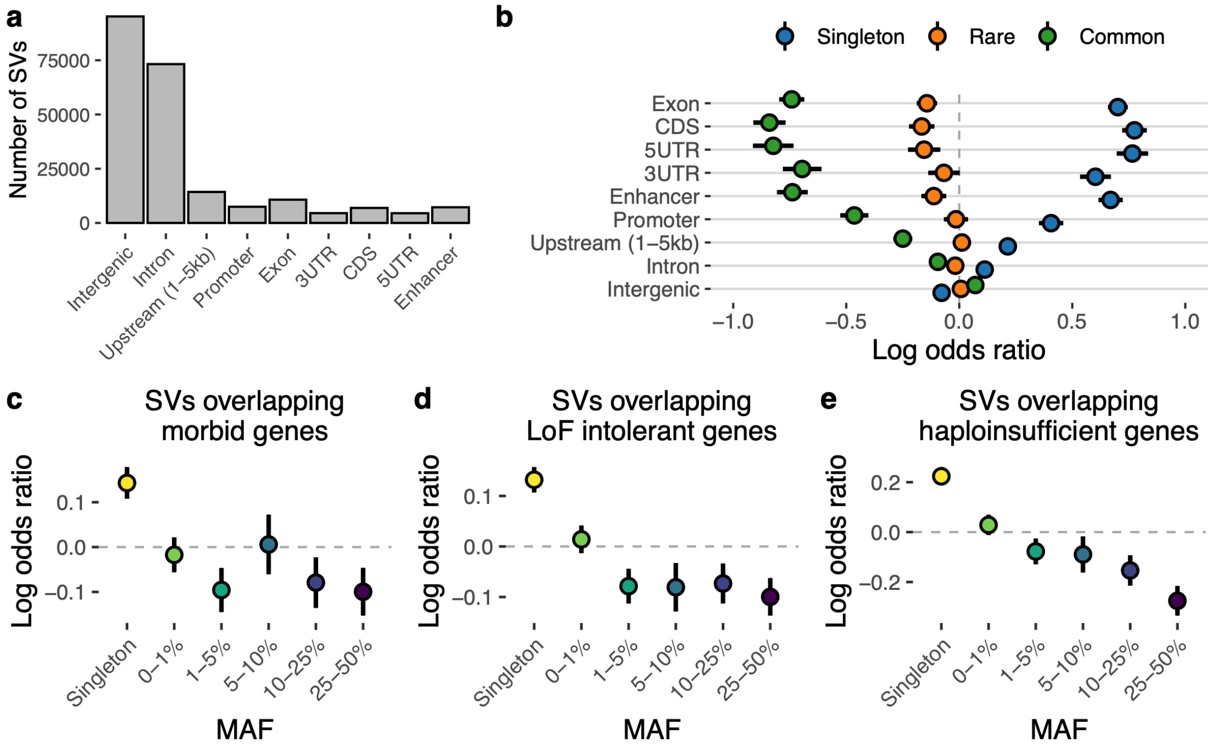


**Supplementary Figure S6: Functional context and evolutionary constraints. a**, Number of SVs overlapping gene and genomic regions. **b**, Enrichment of SVs overlapping each region stratified by common (MAF>5%), rare (MAF<5%), and singleton. Enrichment of OMIM genes (**c**), LoF intolerant genes (**d**), and Haploinsufficient genes (**e**) overlapping SVs in different frequency stratum. Lines in the enrichment plots indicate Wald confidence intervals of the relative odds.

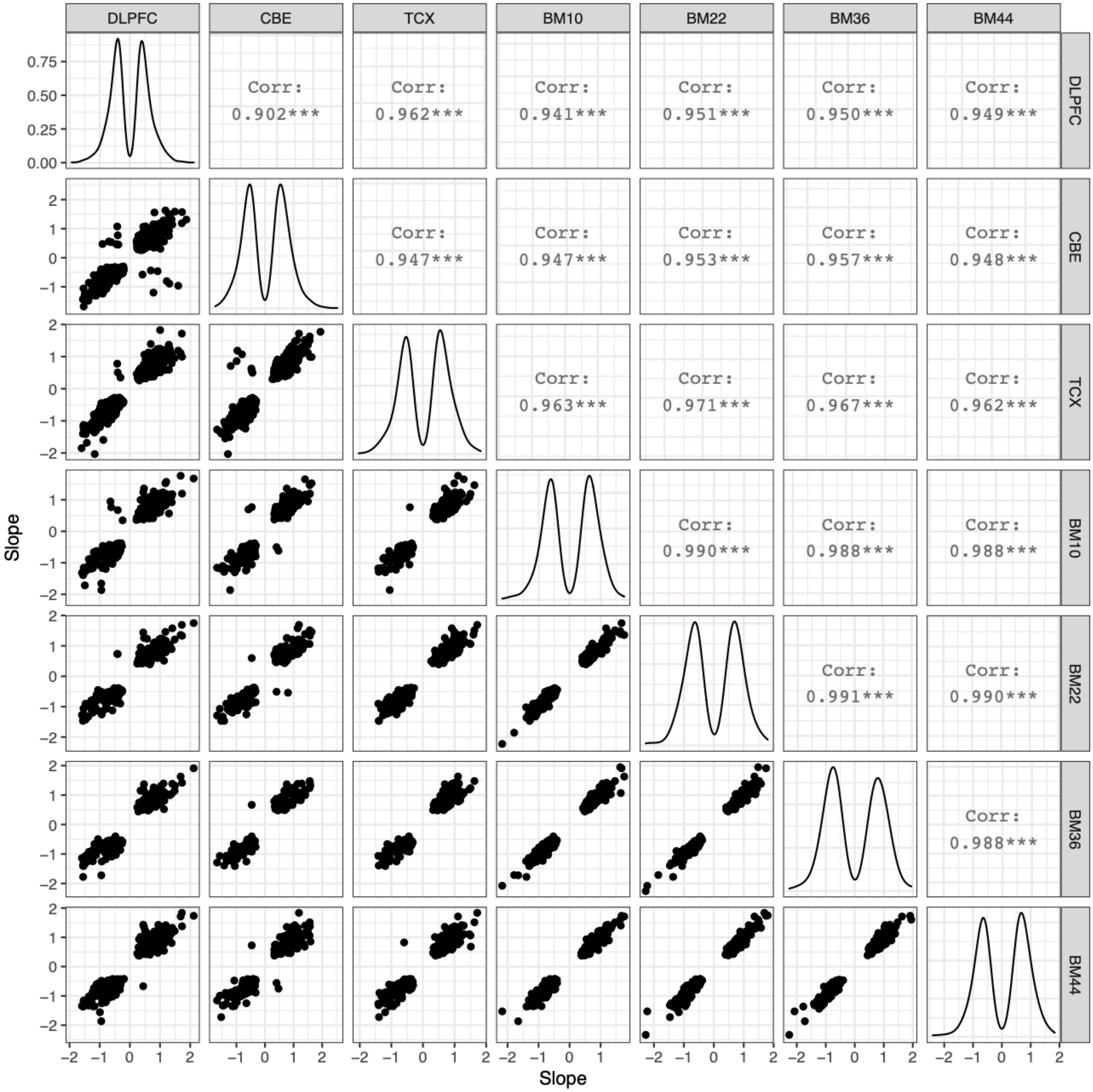


**Supplementary Figure S7: Correlation of direction of effects of SV-eQTLs across brain regions.** Pairwise comparison of SV-eQTLs slopes for each brain region. Only significant SV-eQTLs (FDR < 0.05) were considered for comparison. Plots in the lower triangular grid show scatter plots of slopes. Pearson's correlations are shown in the upper triangular grid. Plots in the diagonal shows the slope distributions.

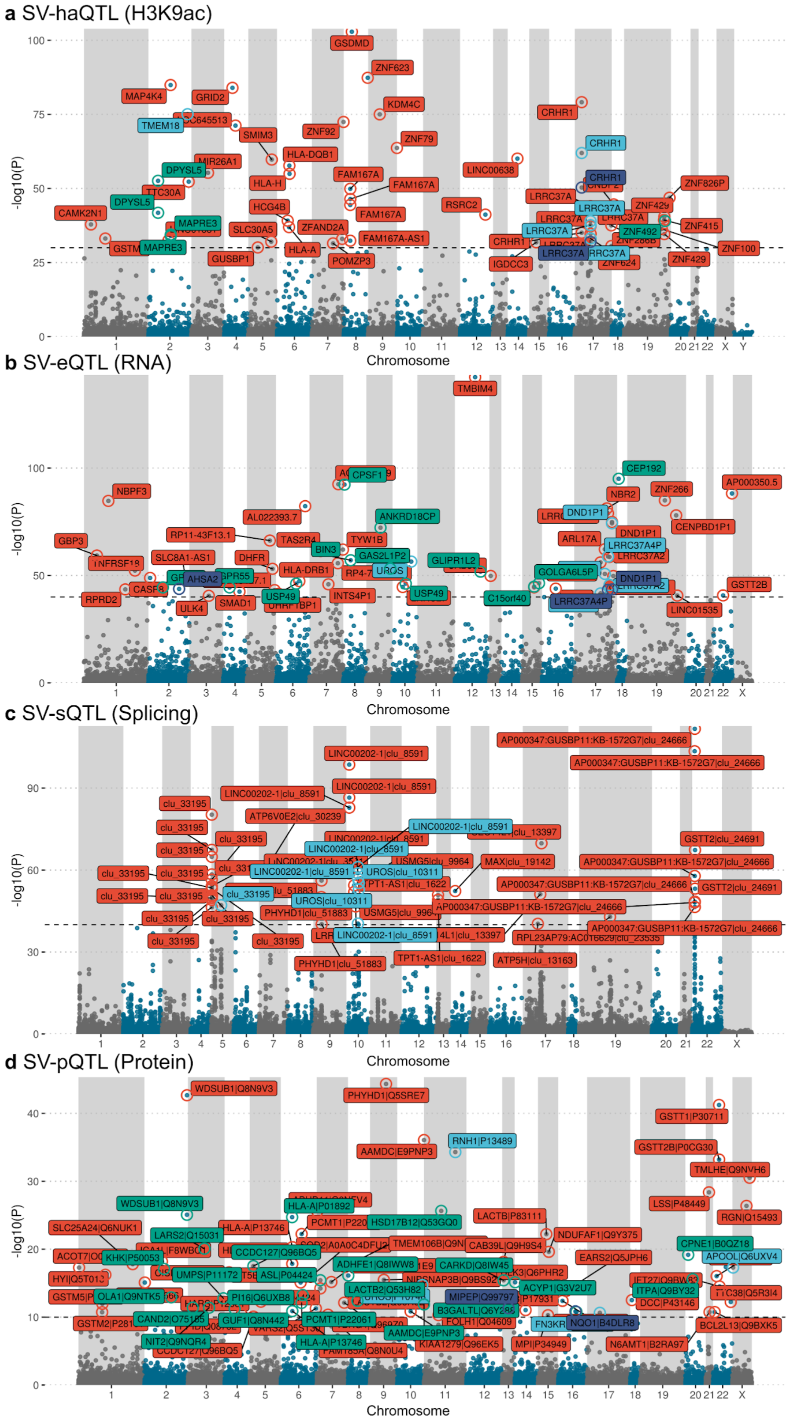


**Supplementary Figure S8: SV-xQTL top hits.** Manhattan plots showing the top SV-xQTLs measured in ROS/MAP. Colored labels represent each SV class. **a**, SV-haQTL (H3K9ac), showing labels for associations with -$\log_{10}$(*P*-value)>30. **b**, SV-eQTL, labels for associations with -$\log_{10}$(*P*-value)>40. **c**, SV-sQTL, labels for associations with -$\log_{10}$(*P*-value)>40. **d**, SV-pQTL, labels for associations with -$\log_{10}$(*P*-value)>10.

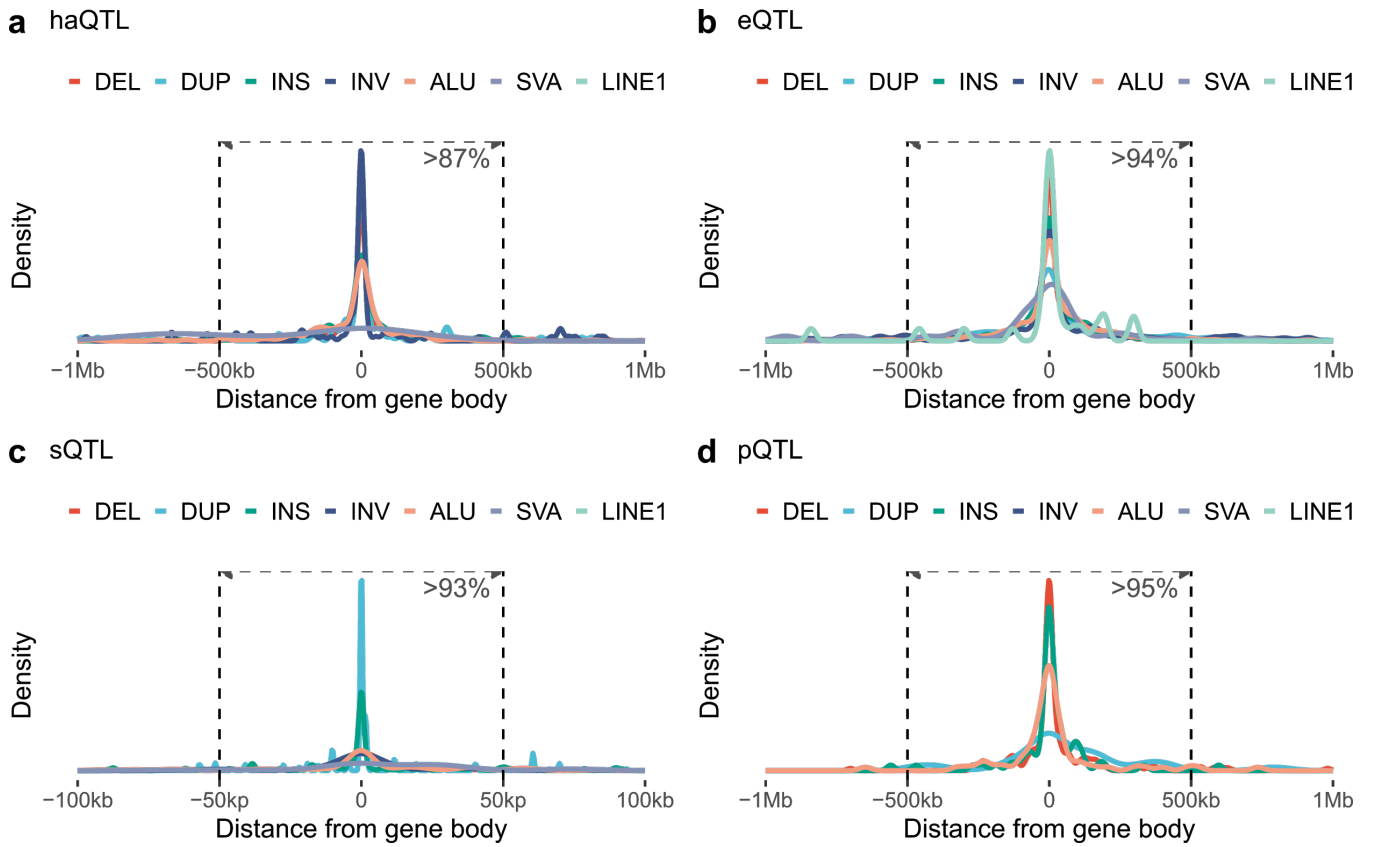


**Supplementary Figure S9: SV distance to the gene body (SV-xQTL).** Distribution of significantly associated SVs by their distance to the gene body (closest breakpoint) measured for each phenotype in ROS/MAP. **a**, SV-haQTLs. **b**, SV-eQTL. **c**, SV-sQTL. **d**, SV-pQTL. For SV-haQTLs the distance of the gene was considered based on the closest gene of the associated peak, while for the other phenotypes was considered the distance to the associated gene.

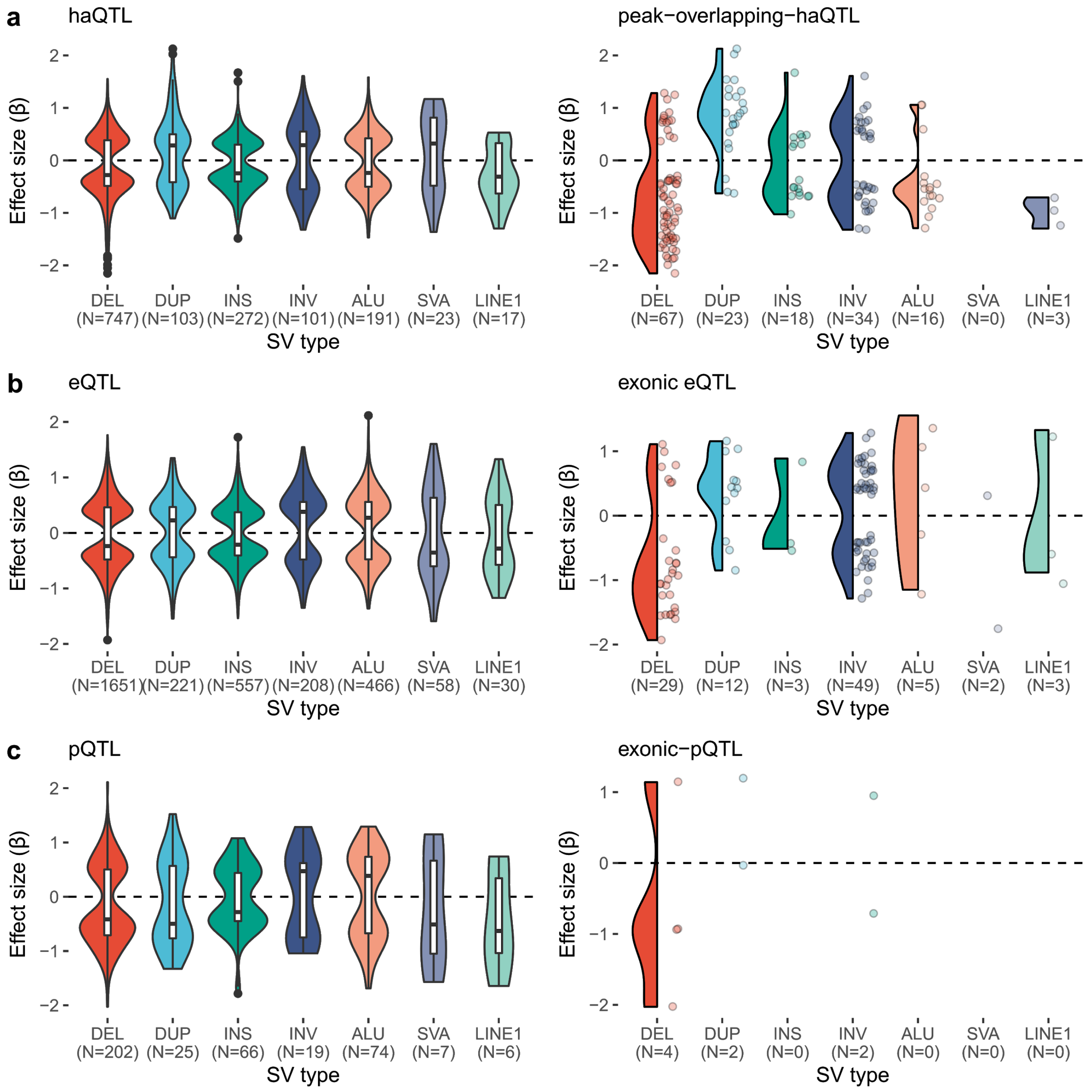


**Supplementary Figure S10: SV-xQTL effect sizes.** Distribution of effect sizes for all SVx-QTLs by SV class. Plots on the left show results for all associated SVs, plots on the right show results only for SVs overlapping either the associated histone peak (SV-haQTL, **a**), or exonic regions of the associated gene (SV-eQTL on **b** and SV-pQTL on **c**).

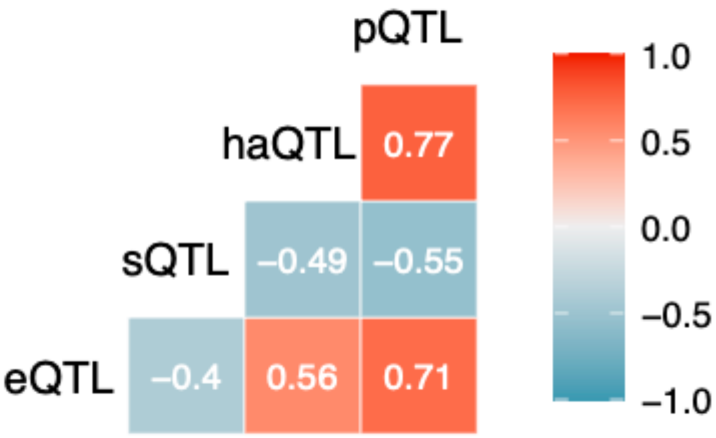


**Supplementary Figure S11: Correlation between SV-xQTL effects.** Pairwise Pearson correlation of effect sizes between each phenotype association.

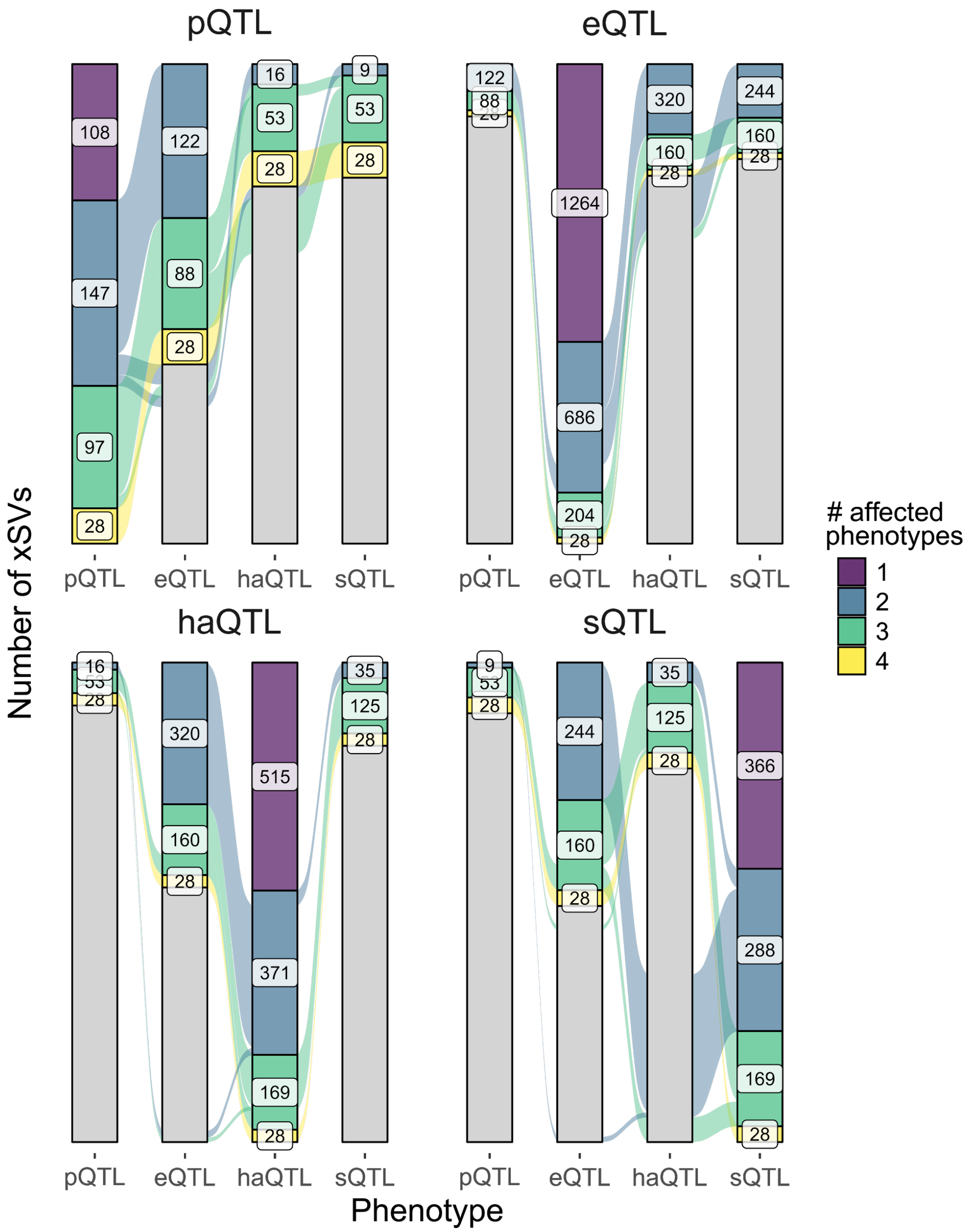


**Supplementary Figure S12: SVs affecting multiple phenotypes in the regulatory cascade.** Each plot subgroup shows the total number of SVs associated with each phenotype and numbers of SVs in each set also associated with different molecular phenotypes. Each color represents SVs affecting different levels (1 to 4) at the same time.

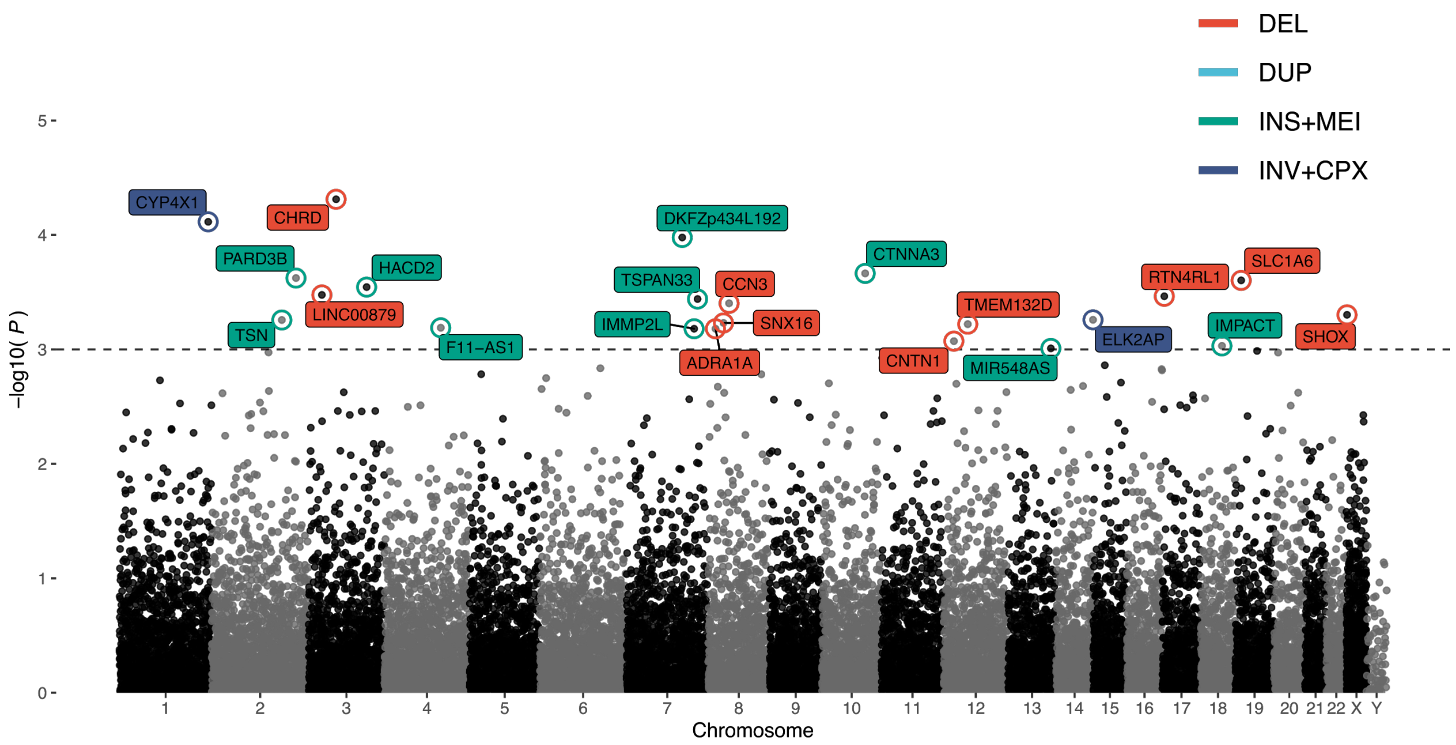


**Supplementary Figure S13: Alzheimer's disease SV associations.** Association tests were run using 22,007 SVs with MAF > 1% SVs identified across ROS/MAP, Mayo Clinic, and MSBB. The y-axis shows the nominal *P*-values given by a logistic regression fitted using 539 AD cases and 368 controls and adjusting for sex, study, and the first three ancestry principal components. Dashed-line represents a suggestive threshold of $-\log_{10}(P\text{-value}) > 3$. Labels indicate the closest gene of each SV. Different colors represent different SV classes.

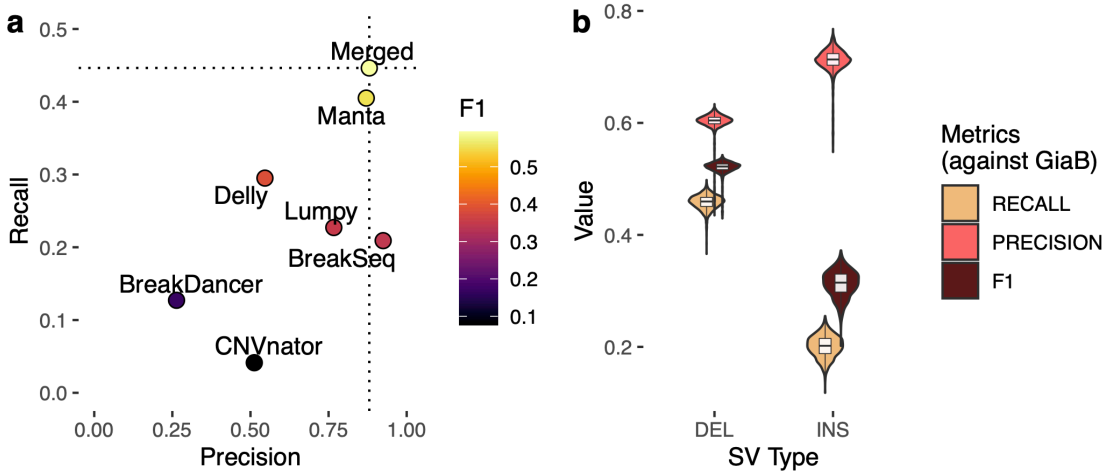


**Supplementary Figure S14: Quality assessment of variant calling.** *In silico* benchmarking and validation. **a**, Benchmarking of individual SV discovery tools and combined tools ("Merged") for the sample HG002 evaluated against the Genome in a Bottle v0.6 Tier 1 using *truvari*. "Merged" strategy was defined by the best F1-score after testing all possible combinations of tools (for insertions and deletions separately). The same merging criteria was applied for all samples in AMP-AD. **b**, Benchmarking results of all AMP-AD samples evaluated against the Genome in a Bottle v0.6 Tier 1 using *truvari*.

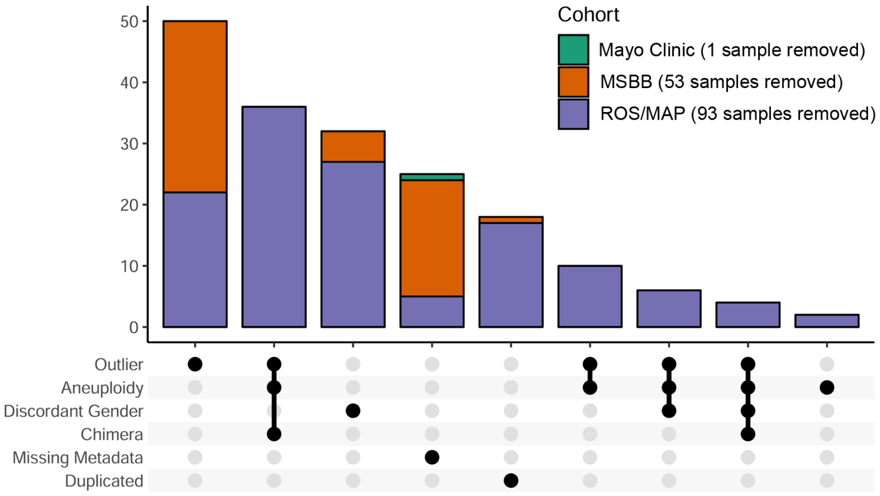


**Supplementary Figure S15: Samples removed.** Upset plot showing the number of samples removed in each cohort according to each QC criteria.

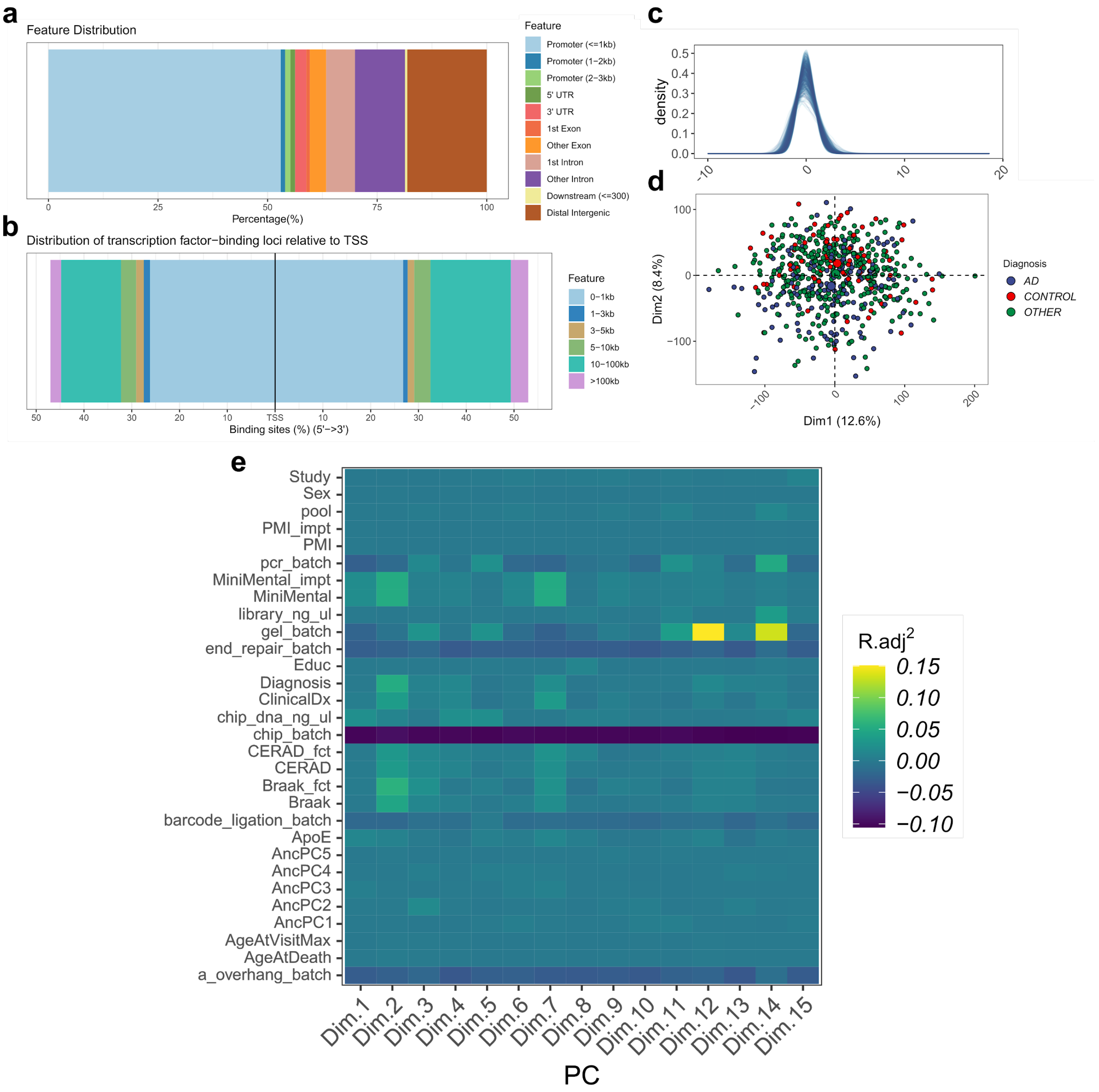


**Supplementary Figure S16: ChIP-seq H3K9ac QC. a**, Genomic feature annotation of H3K9ac peaks and **b**, distribution of transcription factor binding loci relative to TSS, measured using ChIPseeker R package. **c**, Density distribution of H3K9ac estimates, each line represents one sample. **d**, PCA plot representing sample distribution, colored by ROS/MAP diagnosis status (AD = Alzheimer's Disease). **e**, Heatmap of adjusted $R^2$ values measured using linear regression between 15 first PCs against technical and biological covariates.

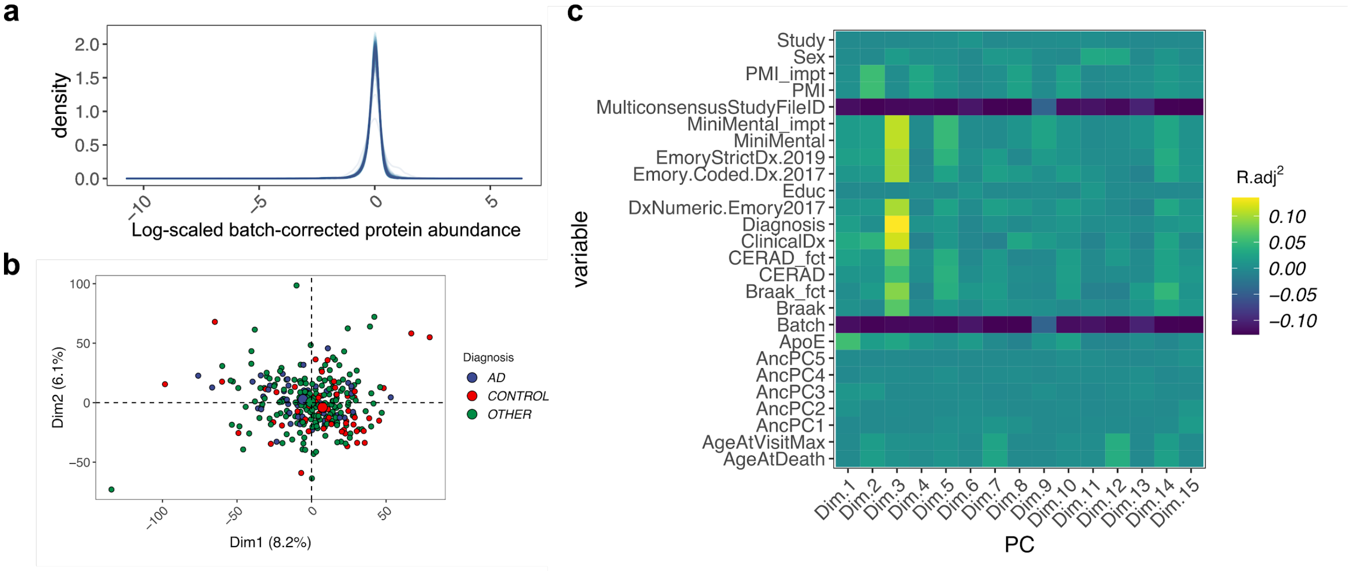


**Supplementary Figure S17: TMT proteomics QC. a**, Density distribution of protein abundances. Values are given in log-scale after batch correction. Each line represents one sample. **b**, PCA plot representing sample distribution, colored by ROS/MAP diagnosis status (AD = Alzheimer's Disease). **c**, Heatmap of adjusted $R^2$ values measured using linear regression between 15 first PCs against technical and biological covariates.

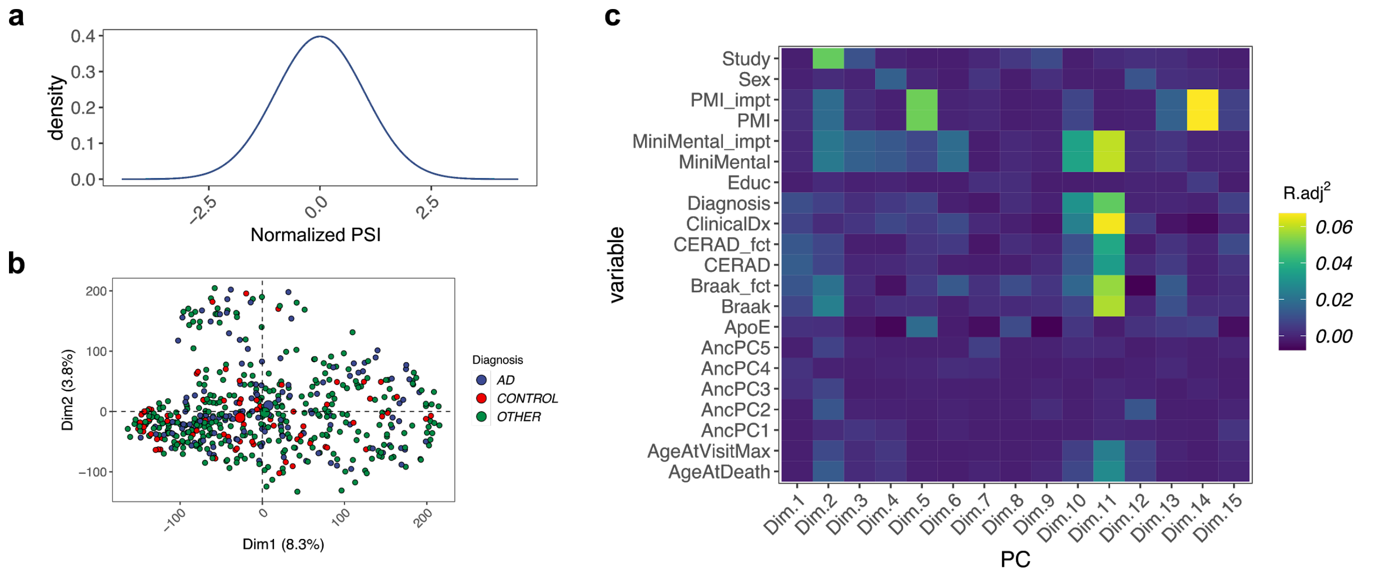


**Supplementary Figure S18: Splicing QC. a**, Density distribution of percent spliced-in (PSI) per sample. Values are standardized across individuals for each intron and quantile normalized across introns. **b**, PCA plot representing sample distribution, colored by ROS/MAP diagnosis status (AD = Alzheimer's Disease). **c**, Heatmap of adjusted $R^2$ values measured using linear regression between 15 first PCs against technical and biological covariates.

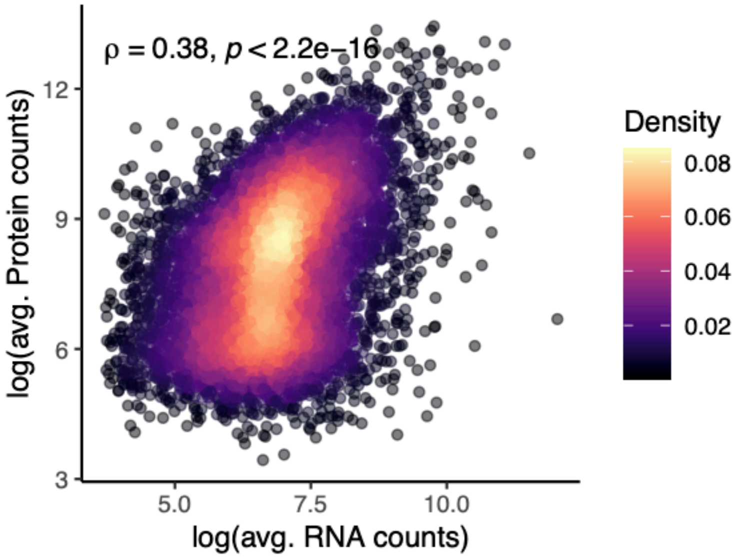


**Supplementary Figure S19: Correlation between RNA and protein expression.** Scatter plot showing expression values for 7,546 genes in 112 samples measured at RNA (x-axis) and protein levels (y-axis). Values are shown as log normalized counts. Spearman rho correlation and *P*-value are shown.

**Supplementary Table S1 - Sample quality control**

| | | ROS/MAP | | | MSBB | | | Mayo Clinic | | |
|---|---|---|---|---|---|---|---|---|---|---|
| **Analysis stage** | **Threshold** | **Eligible samples** | **Sample failures** | **Fail rate** | **Eligible samples** | **Sample failures** | **Fail rate** | **Eligible samples** | **Sample failures** | **Fail rate** |
| *Pre-SV discovery filters* | | | | | | | | | | |
| **All pre-SV discovery metrics** | | **1200** | **71** | **0.059** | **354** | **25** | **0.071** | **350** | **1** | **0.003** |
| Missing metadata information | . | 1200 | 5 | 0.004 | 354 | 19 | 0.054 | 350 | 1 | 0.003 |
| Duplicated IDs | . | 1196 | 17 | 0.014 | 354 | 1 | 0.003 | 350 | 0 | 0 |
| Discordant reported & inferred sex | . | 1178 | 30 | 0.025 | 353 | 5 | 0.014 | 350 | 0 | 0 |
| Pairwise alignment rate | >0.95 | 1178 | 0 | 0 | 353 | 0 | 0 | 350 | 0 | 0 |
| Chimera rate | <0.04 | 1178 | 13 | 0.011 | 353 | 0 | 0 | 350 | 0 | 0 |
| Adapter rate | <0.07 | 1178 | 0 | 0 | 353 | 0 | 0 | 350 | 0 | 0 |
| Read length | >125 | 1178 | 0 | 0 | 353 | 0 | 0 | 350 | 0 | 0 |
| Coverage | >0.2 | 1178 | 0 | 0 | 353 | 0 | 0 | 350 | 0 | 0 |
| Autosomal ploidy spread | <1 | 1178 | 22 | 0.019 | 353 | 0 | 0 | 350 | 0 | 0 |
| *Post-SV discovery filters* | | | | | | | | | | |
| **All post-SV discovery metrics** | | **1178** | **5** | **0.004** | **333** | **28** | **0.084** | **349** | **0** | **0** |
| Failed run SV tool | . | 1178 | | | 333 | 0 | | 349 | 0 | |
| Outlier SV number | 3*IQR | 1178 | 5 | | 333 | 28 | | 349 | 0 | |
| *Final sample set* | | | | | | | | | | |
| **All QC metrics** | | **1200** | **94** | **0.078** | **354** | **49** | **0.138** | **350** | **1** | **0.003** |

**Supplementary Table S2 - Results for SV discovery in HG002 using different tools**

| Tool | Total SVs | Deletions | Duplications | Insertions | Inversions | Translocations |
|---|---|---|---|---|---|---|
| GIAB | 12,745 | 5,464 | - | 7,281 | - | - |
| Manta | 9,105 | 5,131 | 521 | 2,089 | 288 | 1,076 |
| LUMPY | 11,163 | 4,049 | 588 | - | 1,380 | 5,146 |
| Delly | 13,252 | 7,497 | 1,810 | 118 | 1,004 | 2,823 |
| BreakDancer | 9,421 | 3,685 | 2,710 | 1,496 | 632 | 898 |
| BreakSeq | 2,883 | 2,686 | - | 197 | - | - |
| CNVnator | 4,140 | 3,198 | 942 | - | - | - |

**Supplementary Table S3 - Benchmarking results**

| Strategy | SV class | Id (figure) | Recall | Precision | F1 | Desc |
|---|---|---|---|---|---|---|
| 5 | DEL | 2 | 0.7461 | 0.919 | 0.8236 | Manta + At least other 2 |
| 1 | INS | 11 | 0.2161 | 0.9461 | 0.3518 | Manta + BreakSeq2 |

**Supplementary Table S4 - Samples selected for SV validation**

| | Sample (WGS id) | |
|---|---|---|
| | SM-CJEK6 | SM-CJK3B |
| RNA-seq | TRUE | TRUE |
| Proteins | TRUE | TRUE |
| H3K9ac | TRUE | TRUE |
| Study | ROS | MAP |
| Sex | F | M |
| PMI | 4.5 | 7.75 |
| APOE | 33 | 33 |
| Age at death | 84 | 89 |
| BRAAK score | 2 | 3 |
| CERAD score | 3 | 4 |
| Ancestry | EUR | EUR |
| Greedy rank (raw SVs) | 15 | 39 |
| Greedy rank (post-genotyping) | 4 | 60 |
| topN rank (raw SVs) | 40 | 114 |
| topN rank (post-genotyping) | 32 | 230 |

**Supplementary Table S5 - Long read sequencing metrics**

| | Sample | |
|---|---|---|
| | **SM-CJEK6** | **SM-CJK3B** |
| Mean read length | 14,837.70 | 13,638.90 |
| Mean read quality | 40 | 40 |
| Median read length | 14,944 | 13,937 |
| Median read quality | 40 | 40 |
| Number of reads | 3,302,482 | 3,705,283 |
| Read length N50 | 18,348 | 16,564 |
| Total bases | 49,001,302,549 | 50,536,059,165 |
